## Supplementary material for "Effects of trust, risk perception, and health behavior on COVID-19 disease burden: Evidence from a multi-state US survey": S1. English survey

COVID survey for rural states - FINAL

Start of Block: Consent Form

**RESEARCH PARTICIPANT INFORMATION AND CONSENT FORM**
 

**What is the purpose of this study?**
We invite you to take an online survey about the impacts of the coronavirus disease 2019 pandemic (COVID-19) on U.S. residents. The information you provide will help researchers at the University of Idaho and public health officials take action to protect communities from COVID-19. Please take the survey if you are an adult 18 years of age or older. 
 
**What will I do in this study?** 
You will fill out a survey that should take about 20 minutes to complete. You may save your responses at any time and return to the survey at a later time/date.  
 
**Are there any risks to me for participating in the research?**  
Participation in this survey is voluntary. There are no foreseen risks to participation. You may stop this survey at any time or skip a question if you are uncomfortable answering it. We will keep your information anonymous and confidential at all times.
 
**How will I be compensated for my participation?**
As a token of appreciation for your participation, please accept the $1 gift we sent you with the invitation letter.    
 
**Who should I contact for questions or concerns?**
If you have any comments, questions, or concerns about the survey, please contact Ben Ridenhour by or phone: 208-885-8607. 
 
**Statement of consent:** 
By continuing with this survey I am indicating that I am 18 years of age or older and I consent to participate in this research.

| Page Break |
| --- |

End of Block: Consent Form

Start of Block: Risk Perception and Behavioral Responses

The following questions are about the coronavirus disease 2019 pandemic (abbreviated as COVID-19). For the purposes of this survey, we will reference the disease as “COVID-19”. 
Please choose only one answer unless the question says choose all that apply.

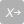

1
Since March 2020, many states have ordered shutdowns in schools, businesses, work, etc due to COVID-19. To what extent have you limited face-to-face interactions with others outside of your household during these shutdowns?

- By a lot (1)
- Some (2)
- By very little (3)

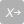

2 How worried are you about each of the following for getting sick with COVID-19?

|  | Not worried at all (1) | Slightly worried (2) | Moderately worried (3) | Very worried (4) |
| --- | --- | --- | --- | --- |
| Yourself (2_1) |  |  |  |  |
| Your family (2_2) |  |  |  |  |
| Others in your community (2_3) |  |  |  |  |

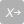

3 How much of a threat do you believe the COVID-19 pandemic is for each of the following?

|  | Not a threat (1) | Minor threat (2) | Major threat (3) |
| --- | --- | --- | --- |
| Your personal health (3_1) |  |  |  |
| Your personal financial situation (3_2) |  |  |  |
| Your local education system (3_3) |  |  |  |
| The economy of your community (3_4) |  |  |  |
| The health of your community (3_5) |  |  |  |
| The health of the US population as a whole (3_6) |  |  |  |
| The US economy (3_7) |  |  |  |

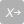

4 Think about the 12 months PRIOR to when you first heard about COVID-19. How often did you engage in the below activities during that pre-COVID period?

|  | Never (1) | Occasionally (2) | Often (3) | Very often (4) |
| --- | --- | --- | --- | --- |
| Gather indoors with close friends or family who do not live in your home (4_1) |  |  |  |  |
| Gather indoors with a large group of 20-30 friends or co-workers (4_2) |  |  |  |  |
| Eat inside at a restaurant (4_3) |  |  |  |  |
| Attend indoor church or faith services (4_4) |  |  |  |  |
| Go shopping in your town (4_5) |  |  |  |  |
| Go to an appointment for half an hour or longer (e.g. haircut, dental cleaning) (4_6) |  |  |  |  |
| Participate in community activities or events like sports, fairs, and festivals, or let your kids do so (4_7) |  |  |  |  |

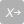

5 Think about your COVID-19 situation right now. Given this situation, which of the following activities do you engage in currently?

|  | Yes (1) | No (2) |
| --- | --- | --- |
| Gather indoors with close friends or family who do not live in your home (5_1) |  |  |
| Gather indoors with a large group of 20-30 friends or co-workers (5_2) |  |  |
| Eat inside at a restaurant (5_3) |  |  |
| Attend indoor church or faith services (5_4) |  |  |
| Go shopping in your town (5_5) |  |  |
| Go to an appointment for half an hour or longer (e.g. haircut, dental cleaning) (5_6) |  |  |
| Participate in community activities or events like sports, fairs, and festivals, or let your kids do so (5_7) |  |  |
| Wear a face mask in public (5_8) |  |  |

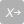

6
Suppose that new cases of COVID-19 are increasing in your state but no new cases are reported in your community. Which of the following activities would you feel comfortable doing?

|  | Yes (1) | No (2) |
| --- | --- | --- |
| Gather indoors with close friends or family who do not live in your home (6_1) |  |  |
| Gather indoors with a large group of 20-30 friends or co-workers (6_2) |  |  |
| Eat inside at a restaurant (6_3) |  |  |
| Attend indoor church or faith services (6_4) |  |  |
| Go shopping in your town (6_5) |  |  |
| Go to an appointment for half an hour or longer (e.g. haircut, dental cleaning) (6_6) |  |  |
| Participate in community activities or events like sports, fairs, and festivals, or let your kids do so (6_7) |  |  |
| Wear a face mask in public (6_8) |  |  |

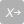

7 Suppose that new cases of COVID-19 are increasing in your community but no cases were reported among people you are around regularly. Which of the following activities would you feel comfortable doing?

|  | Yes (1) | No (2) |
| --- | --- | --- |
| Gather indoors with close friends or family who do not live in your home (7_1) |  |  |
| Gather indoors with a large group of 20-30 friends or co-workers (7_2) |  |  |
| Eat inside at a restaurant (7_3) |  |  |
| Attend indoor church or faith services (7_4) |  |  |
| Go shopping in your town (7_5) |  |  |
| Go to an appointment for half an hour or longer (e.g. haircut, dental cleaning) (7_6) |  |  |
| Participate in community activities or events like sports, fairs, and festivals, or let your kids do so (7_7) |  |  |
| Wear a face mask in public (7_8) |  |  |

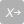

8 Suppose that new cases of COVID-19 are increasing in your community and there were multiple cases reported among people you are around regularly. Which of the following activities would you feel comfortable doing?

|  | Yes (1) | No (2) |
| --- | --- | --- |
| Gather indoors with close friends or family who do not live in your home (8_1) |  |  |
| Gather indoors with a large group of 20-30 friends or co-workers (8_2) |  |  |
| Eat inside at a restaurant (8_3) |  |  |
| Attend indoor church or faith services (8_4) |  |  |
| Go shopping in your town (8_5) |  |  |
| Go to an appointment for half an hour or longer (e.g. haircut, dental cleaning) (8_6) |  |  |
| Participate in community activities or events like sports, fairs, and festivals, or let your kids do so (8_7) |  |  |
| Wear a face mask in public (8_8) |  |  |

End of Block: Risk Perception and Behavioral Responses

Start of Block: Social influence I

You are about to be asked 3 questions about activities in daily life. To answer these questions, suppose that new cases of COVID-19 are increasing in your state, but there are no new cases in your community.

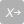

SI1: 9
Suppose your town has no mask-wearing requirement. You go to a store. Would you wear a mask if...

- No other customer wears a mask (1)
- Some customers (1 in 4) wear a mask (2)
- Half of the customers (2 in 4) wear a mask (3)
- Most customers (3 in 4) wear a mask (4)
- Every customer wears a mask (5)
- I would NOT wear a mask even if all customers wear a mask (6)

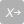

SI1: 10 Suppose a friend is having an indoor gathering. Would you decline (or say “no”) to an invitation to this gathering if...

- No one else declined (1)
- Some guests (1 in 4) declined (2)
- Half of the guests (2 in 4) declined (3)
- Most guests (3 in 4) declined (4)
- All guests declined (5)
- I would ACCEPT the invitation even if everyone else declined (6)

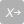

SI1: 11 Suppose a close relative is having an indoor birthday party of your family and close friends. Would you decline (or say “no”) to an invitation to this party if...

- No one else declined (1)
- Some guests (1 in 4) declined (2)
- Half of the guests (2 in 4) declined (3)
- Most guests (3 in 4) declined (4)
- All guests declined (5)
- I would ACCEPT the invitation even if everyone else declined (6)

End of Block: Social influence I

Start of Block: Social Influence II

You are about to be asked 3 questions about activities in daily life. To answer these questions, suppose that new cases of COVID-19 are increasing in your community, but there are no cases among people you interact with regularly.

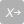

SI2: 9 Suppose your town has no mask-wearing requirement. You go to a store. Would you wear a mask if...

- No other customer wears a mask (1)
- Some customers (1 in 4) wear a mask (2)
- Half of the customers (2 in 4) wear a mask (3)
- Most customers (3 in 4) wear a mask (4)
- Every customer wears a mask (5)
- I would NOT wear a mask even if all customers wear a mask (6)

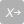

SI2: 10 Suppose a friend is having an indoor gathering. Would you decline (or say “no”) to an invitation to this gathering if...

- No one else declined (1)
- Some guests (1 in 4) declined (2)
- Half of the guests (2 in 4) declined (3)
- Most guests (3 in 4) declined (4)
- All guests declined (5)
- I would ACCEPT the invitation even if everyone else declined (6)

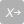

SI2: 11 Suppose a close relative is having an indoor birthday party of your family and close friends. Would you decline (or say “no”) to an invitation to this party if...

- No one else declined (1)
- Some guests (1 in 4) declined (2)
- Half of the guests (2 in 4) declined (3)
- Most guests (3 in 4) declined (4)
- All guests declined (5)
- I would ACCEPT the invitation even if everyone else declined (6)

End of Block: Social Influence II

Start of Block: Social Influence III

You are about to be asked 3 questions about activities in daily life. To answer these questions, suppose that new cases of COVID-19 are increasing in your community AND there are multiple cases among people you interact with regularly.

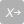

SI3: 9 Suppose your town has no mask-wearing requirement. You go to a store. Would you wear a mask if...

- No other customer wears a mask (1)
- Some customers (1 in 4) wear a mask (2)
- Half of the customers (2 in 4) wear a mask (3)
- Most customers (3 in 4) wear a mask (4)
- Every customer wears a mask (5)
- I would NOT wear a mask even if all customers wear a mask (6)

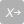

SI3: 10 Suppose a friend is having an indoor gathering. Would you decline (or say “no”) to an invitation to this gathering if...

- No one else declined (1)
- Some guests (1 in 4) declined (2)
- Half of the guests (2 in 4) declined (3)
- Most guests (3 in 4) declined (4)
- All guests declined (5)
- I would ACCEPT the invitation even if everyone else declined (6)

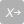

SI3: 11 Suppose a close relative is having an indoor birthday party of your family and close friends. Would you decline (or say “no”) to an invitation to this party if...

- No one else declined (1)
- Some guests (1 in 4) declined (2)
- Half of the guests (2 in 4) declined (3)
- Most guests (3 in 4) declined (4)
- All guests declined (5)
- I would ACCEPT the invitation even if everyone else declined (6)

End of Block: Social Influence III

Start of Block: Social Influence Cont.

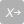

12 Which one of the below statements best describes your view about COVID-19?

- COVID-19 is not real (1)
- COVID-19 is real but not serious (2)
- COVID-19 is real and serious (3)

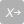

13 How strongly do you agree or disagree that shutting down schools and businesses to reduce the spread of COVID-19 has come at too great an economic cost?

- Strongly agree (1)
- Somewhat agree (2)
- Somewhat disagree (3)
- Strongly disagree (4)

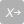

14 About how common is it for your family and close friends to share similar views as yours on the economic cost of school and business shutdowns due to COVID-19? Please give us your best estimate.

- Very rare (fewer than 1 in 4 people) (1)
- Rare (1 in 4 people) (2)
- About even (2 in 4 people) (3)
- Common (3 in 4 people) (4)
- Very common (more than 3 in 4 people) (5)

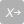

15 About how common is it for people in your community to share similar views as yours on the economic cost of school and business shutdowns due to COVID-19? Please give us your best estimate.

- Very rare (fewer than 1 in 4 people) (1)
- Rare (1 in 4 people) (2)
- About even (2 in 4 people) (3)
- Common (3 in 4 people) (4)
- Very common (more than 3 in 4 people) (5)

End of Block: Social Influence Cont.

Start of Block: Work and Economy

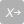

16 What was your employment status at the beginning of 2020, prior to the COVID-19 pandemic?

- I was employed for wages full time (1)
- I was employed for wages part time (2)
- I was self-employed (3)
- I was unemployed and looking for work (4)
- I was unemployed and not looking for work (e.g. retired, disabled) (5)

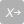

17 Did you experience any of the following because of the COVID-19 pandemic?

|  | Yes (1) | No (2) |
| --- | --- | --- |
| Worked fewer hours (17_1) |  |  |
| Was laid off (17_2) |  |  |
| Took a pay cut (17_3) |  |  |
| Had to take unpaid time off (17_4) |  |  |
| None of the above (17_5) |  |  |
| Not applicable (e.g. not employed) (17_6) |  |  |

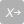

18 How hard is it for you to pay your household bills right now?

- Not hard (1)
- Somewhat hard (2)
- Very hard (3)

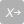

19 If you were to develop mild symptoms of a respiratory infection this week (e.g. cough or fever or shortness of breath), would you...

|  | Yes (1) | No (2) |
| --- | --- | --- |
| Isolate at home for at least 10 days (19_1) |  |  |
| Isolate at home until the symptoms improve (19_2) |  |  |
| Try to get tested for COVID-19 (19_3) |  |  |
| Go to work (19_4) |  |  |
| Continue seeing friends and family (19_5) |  |  |

Skip To: 20 If If you were to develop mild symptoms of a respiratory infection this week (e.g. cough or fever or... = Go to work [ Yes ]

Skip To: End of Block If If you were to develop mild symptoms of a respiratory infection this week (e.g. cough or fever or... = Go to work [ No ]

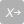

20 If you were to develop mild symptoms of a respiratory infection this week (e.g. cough or fever or shortness of breath), you would go to work because...

|  | Yes (1) | No (2) |
| --- | --- | --- |
| You are not worried about the coronavirus (20_1) |  |  |
| You need to earn money (20_2) |  |  |
| You are afraid of losing your job or having your hours reduced (20_3) |  |  |
| Your work is too important to miss (20_4) |  |  |
| Not applicable to you (e.g. not employed, work from home) (20_5) |  |  |
| Other reasons (please specify) (20_6) |  |  |

End of Block: Work and Economy

Start of Block: Contact Tracing, Vaccines, Trust

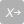

21 Contact tracing is the idea that when someone tests positive for COVID-19, the people they have come in close contact with should be notified and tested. If a contact tracer called you because you may have been exposed, how likely are you to follow each of the recommendations below?

|  | Very Unlikely (1) | Unlikely (2) | Likely (3) | Very Likely (4) |
| --- | --- | --- | --- | --- |
| Self-isolate at home for 10 days (21_1) |  |  |  |  |
| Get tested for the virus if you develop symptoms (21_2) |  |  |  |  |
| Get tested for the virus even if you do not develop symptoms (21_3) |  |  |  |  |
| Share with the contact tracer the names of people you have been in close contact with recently (21_4) |  |  |  |  |

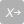

22 If a vaccine against COVID-19 is approved by the FDA (Food and Drug Administration) and becomes available, would you get vaccinated?

- Yes (1)
- No (2)
- Maybe--it depends on the number of new COVID-19 cases at the time (3)
- Maybe--it depends on the side-effects of the vaccine (4)
- Maybe--it depends on both the number of new COVID-19 cases at the time and the side-effects of the vaccine (5)

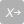

23 Assuming the vaccine against COVID-19 is highly effective and has side-effects similar to the flu vaccine, when would you get vaccinated? 

- I would get vaccinated no matter what (1)
- If new cases of COVID-19 are steady in my state (not going up or down), but there are no new cases in my community (2)
- If new cases of COVID-19 are increasing rapidly in my state, but there are no new cases in my community (3)
- If new cases of COVID-19 are increasing rapidly in my community, but there are no cases among people I interact with regularly (4)
- If new cases of COVID-19 are increasing rapidly in my community, and there are multiple cases among people I interact with regularly (5)
- I would never get vaccinated (6)

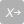

24 Indicate how informed you feel about the following.

|  | Very informed (1) | Somewhat informed (2) | Not at all informed (3) |
| --- | --- | --- | --- |
| Symptoms of COVID-19 (24_1) |  |  |  |
| How to avoid getting COVID-19 (24_2) |  |  |  |
| How to get tested for COVID-19 (24_3) |  |  |  |
| Steps to take if someone in your household gets COVID-19 (24_4) |  |  |  |

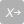

25 Which of the below statements best reflect the status of COVID-19 in your county right now?

- There have been no known new cases of COVID-19 in my county in the past week (1)
- New cases of COVID-19 have gone down in my county in the past week (2)
- New cases of COVID-19 are steady in my county (not going up or down) in the past week (3)
- New cases of COVID-19 have been increasing in my county in the past week (4)
- I don't know (5)

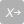

26 How much do you trust the following individuals/groups to provide you with accurate information about COVID-19?

|  | Not at all (1) | A little bit (2) | Some (3) | A lot (4) |
| --- | --- | --- | --- | --- |
| Governor of your state (26_1) |  |  |  |  |
| State/local public health officials (26_2) |  |  |  |  |
| Mayor or county commissioner (26_3) |  |  |  |  |
| Faith leaders (26_4) |  |  |  |  |
| A doctor or health care provider (26_5) |  |  |  |  |
| The Centers for Disease Control and Prevention (CDC) (26_6) |  |  |  |  |
| The World Health Organization (WHO) (26_7) |  |  |  |  |
| University researchers (e.g. epidemiologists) (26_8) |  |  |  |  |
| The Trump Administration (26_9) |  |  |  |  |

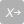

27 How much information do you get from the following sources?

|  | None (1) | A little bit (2) | Some (3) | A lot (4) |
| --- | --- | --- | --- | --- |
| Mainstream Media (Associated Press, New York Times, Wall Street Journal, USA Today, or Washington Post) (27_1) |  |  |  |  |
| Conservative Media (Fox News, Rush Limbaugh, Breitbart News, One America News, or The Drudge Report) (27_2) |  |  |  |  |
| Mainstream Broadcast Media (ABC News, CBS News, NBC News, Univision) (27_3) |  |  |  |  |
| Liberal Media (MSNBC, Vox, Vice, or Huffington Post) (27_4) |  |  |  |  |
| Online News Aggregators (Google News or Yahoo News) (27_5) |  |  |  |  |
| Social Media (Facebook, Twitter, WhatsApp, or YouTube) (27_6) |  |  |  |  |
| The radio (27_7) |  |  |  |  |

End of Block: Contact Tracing, Vaccines, Trust

Start of Block: School; Health

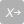

28 Do you have any children under 18 living with you who attend school outside of your house?

- Yes (1)
- No (2)

Skip To: 30 If Do you have any children under 18 living with you who attend school outside of your house? = No

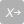

29 Assuming in-person schooling does not close in your community, when would you keep your children home from school?

If new cases of COVID-19 are steady in my state (not going up or down), but there are no new cases in my community (1)

If new cases of COVID-19 are increasing rapidly in my state, but there are no new cases in my community (2)

If new cases of COVID-19 are increasing rapidly in my community, but there are no cases among people I interact with regularly (3)

If new cases of COVID-19 are increasing rapidly in my community, and there are multiple cases among people I interact with regularly (4)

Regardless of the COVID-19 situation, I would send kids to school as long as school is open (5)

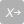

30 Do you have health insurance?

- Yes (1)
- No (2)

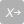

31
The CDC defines the following categories as high-risk categories for COVID-19. Indicate if any of these apply to you. Choose all that apply.

- I am 65 years or older (1)
- I am immune compromised or am taking medication that suppresses my immune system (2)
- I have a health condition (such as cancer, kidney disease, Type 2 diabetes, heart disease, lung disease, obesity, sickle cell disease) (3)
- I am not in a high-risk category (4)

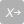

32
Have you had COVID-19?

- Yes--I tested positive (1)
- Probably--For example, I had symptoms but was never tested (2)
- No--I don't think so (3)
- I don't know or I'm not sure (4)
- I prefer not to say (5)

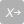

33 How many people you personally know have had COVID-19 or believe they have had it?

- None (1)
- 1 to 5 (2)
- More than 5 (3)
- I don't know or I'm not sure (4)
- I prefer not to say (5)

End of Block: School; Health

Start of Block: Demographics

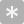

34 How many people currently live in your household, including you? (Write a number)

________________________________________________________________

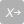

35 Besides yourself and your spouse/partner, do any people 65 or older currently live in your household?

- Yes (1)
- No (2)

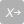

36 Besides yourself, is there anybody in your household that may be at high risk of severe COVID-19?

- Yes (1)
- No (2)
- I prefer not to say (3)

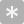

37 In what year were you born? (Write a number)

________________________________________________________________

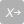

38 Are you Hispanic or Latino?

- Yes (1)
- No (2)

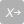

39 Which categories describe you? Choose all that apply.

- American Indian or Alaska Native (1)
- Asian or Asian-American (2)
- Black or African American (3)
- Native Hawaiian or Other Pacific Islander (4)
- White (5)
- Other (please specify) (6) ________________________________________________

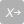

40 What is the highest level of formal education that you have completed?

- Did not graduate high school (1)
- High school diploma or equivalent (GED) (2)
- Some college (3)
- Associate’s Degree (4)
- College graduate or higher (5)

41
What is your occupation?

________________________________________________________________

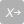

42 What is your religion/faith?

- Evangelical / Nondenominational Christian (1)
- Catholic (2)
- Mainline Protestant Christian (3)
- Latter-Day Saints (4)
- Jewish (5)
- Islam (6)
- Buddhist (7)
- Atheist (8)
- Agnostic (9)
- Other faiths / other religion (please specify) (10) ________________________________________________

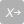

43 How would you describe your political views?

- Very liberal (1)
- Liberal (2)
- Moderate (3)
- Conservative (4)
- Very conservative (5)
- Libertarian (6)
- Non-political (7)
- Other (please specify) (8) ________________________________________________

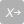

44 What is your marital status?

- Single, never married (1)
- Married or domestic partnership (2)
- Widowed (3)
- Divorced (4)
- Separated (5)

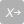

45 In 2019, what was your total household income from all sources before taxes?

- Less than $25,000 (1)
- $25,000 - 49,999 (2)
- $50,000 - 74,999 (3)
- $75,000 or more (4)

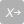

46 What is your gender?

- Male (1)
- Female (2)
- Do not identify as male or female (3)

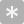

47 What is your zip code?

________________________________________________________________

End of Block: Demographics

Start of Block: Final Comments

48
Thank you for taking the time to fill out our survey! We really appreciate your participation. If you would like to take part in a follow-up interview about your opinions on COVID-19 please provide us with your contact information (phone number or email) below.

________________________________________________________________

49 If you have any additional comments about COVID-19 or this survey, please feel free write them in the space provided below.

________________________________________________________________

________________________________________________________________

________________________________________________________________

________________________________________________________________

________________________________________________________________

End of Block: Final Comments
