## Supplementary material for "Effects of trust, risk perception, and health behavior on COVID-19 disease burden: Evidence from a multi-state US survey": S2. R markdown: Supplemental_Appendix.Rmd

````
---
title: "Supplemental Appendix"
fontsize: 12pt
output:
  pdf_document:
    toc: yes
    toc_depth: 2
    fig_caption: yes
    number_sections: no
  linestretch: 1.5
  header-includes:
    \usepackage{pdflscape}
    \newcommand{\blandscape}{\begin{landscape}}
    \newcommand{\elandscape}{\end{landscape}}
---


```{r setup, include=FALSE}
knitr::opts_chunk$set(echo = TRUE, fig.width = 10, fig.height = 10)
library(lavaan)
library(semPlot)
library(sjPlot)
library(officer)
library(dplyr)
library(plyr)
```


\newpage
## Supplemental Appendix Overview

This supplemental appendix provides full analysis outputs, associated reproducible code, and supportive data. As noted in the associated paper, our analysis describes relationships between social cognitive measures (collected from our COVID-19 survey, conducted in the Fall of 2020) and COVID-19 cases (aggregated by the New York Times - http://github.com/nytimes/covid19).  


**Data Collection and Transformation**

Our structural equation models rely upon the following datasets:

1. COVID-19 daily case data, at a county level.  We use aggregate case data that is assembled by the New York Times, and is exposed via their github.com repository (http://github.com/nytimes/covid19).  This dataset is within our /data folder, named "COVID_data.csv".

2. COVID-19 survey data.  We conducted a COVID-19 health perceptions survey in the Fall of 2020 (n = 1035). This de-identified survey dataset is within our /data folder, named "COVID_survey.csv". 

From a transformation perspective, several additional fields are generated:

1. Data is reduced to only cases for Idaho, Texas and Vermont;
2. COVID-19 cases per county are grouped based on 1) cumulative cases between June 1, 2020 and April 30th, 2021, as well as 2) cumulative cases 15 days before and after survey completion. 
3. In order to assign rurality, survey responses were grouped into urban and rural classifications, based on the United States Department of Agriculture's Rural/Urban Commuting Areas (RUCA)
\newpage

**Conceptual Models Overview**

As note in our associated paper, we evaluated 6 (six) models.  Please note the following variable naming conventions: 1) 'Health Risk' and 'Economic Risk' refer to health risk perception and economic risk perception, respectfully. 2) "Behavior" refers to both current behavior (at the time of survey completion) as well as behavioral intention. Shorter terms are used for brevity in diagram notation.

**Model Set A:** Behavioral intentions as an influence on disease burden. Here we consider the disease burden as cumulative cases across a large window of time (June 2020 to April 2021) to capture a complete view of disease impact.  

* Model 1A: Trust effects on economic risk perception and health risk perception, to behavioral intention, and then to disease burden.
* Model 2A: Economic risk perception and health risk perception effects on trust, which impact behavioral intention, and then disease burden.
* Model 3A: Trust, economic risk perception and health risk perception - all impacting behavioral intention directly, and then to disease burden.

**Model Set B:** Disease burden as an influence on behavioral intention. Here we consider a snapshot of the cases in the two weeks prior to the survey response as a measure of the local disease burden and consider its impact on the expressed behavioral intention.

* Model 1B: Trust effects on economic risk perception and health risk perception, and then behavioral intention.
* Model 2B: Economic risk perception and health risk perception effects on trust, which impact behavioral intention.
* Model 3B: Trust, economic risk perception and health risk perception - all impacting behavioral intention directly.

\newpage

```{r data_setup, echo=FALSE, message=FALSE, warning=FALSE}
#SurveyResults, deidentified with geography
fulldata <- read.csv(file="./data/COVID_survey_original.csv")
#Cumulative Daily Disease data by county
coviddata<-read.csv("./data/us_counties_covid.csv")
coviddata<-dplyr::rename(coviddata, FIPS = fips)[,c("date","FIPS","cases")]
coviddata<-semi_join(coviddata,fulldata)
US_census<-read.csv("./data/US_population_county_ACS_2018_5yr.csv")
US_census$FIPS<-substr(US_census$id, nchar(US_census$id)-4, nchar(US_census$id))
US_census %>% dplyr::rename(CountyPop = Estimate..Total)->US_census

fulldata %>% select(c("RegionAbbr", "Urb_Rur", "FIPS", "RecordedDate", "ExternalReference", "Q26_1", "Q26_2", "Q26_3", "Q26_5", "Q26_6", "Q26_7", "Q26_8", "Q46", "Q43", "Q39", "Q37") | contains(c("Q2_", "Q3_","Q4_", "Q5_", "Q6_","Q7_","Q8_"))) -> used

used <- used[used$Q37 != 1902 | is.na(used$Q37),]    
  
used %>% mutate(age65plusbool = as.integer(2020 - Q37 > 64)) %>% 
  mutate(femalebool = as.integer(Q46 == 2))  %>%
  mutate(whitebool = as.integer(Q39 == 5)) %>% 
  mutate(ruralbool = as.integer(Urb_Rur == "R"))-> used

used$RecordedDate <- as.Date(used$RecordedDate, "%m/%d/%y %H:%M")
used<-merge(used,US_census[,c("FIPS","CountyPop")], by="FIPS") #Add county population
used<-merge(used,(coviddata %>% filter(date == "2021-04-30"))[,c("FIPS","cases")], all.x=T) #enddate for cases
used %>% dplyr::rename(casesEND = cases)%>%mutate(date = RecordedDate) -> used #getting casestodate
used<-merge(used,coviddata, all.x=T) 
used %>% dplyr::rename(cumcasestodate = cases)%>%mutate(date = NULL, cumcasesper100 = (casesEND)*100/CountyPop, casetodateper100 = cumcasestodate*100/CountyPop) -> used 
polVec <- factor(c("Liberal","Liberal", "Moderate", "Conservative", "Conservative", "Libertarian", "Nonpolitical", "Other"),
                 levels = c("Moderate","Liberal","Conservative","Libertarian","Nonpolitical","Other"))
used["polVec"] <- polVec[used$Q43]
newframe <- model.matrix(~ polVec + ExternalReference, data = used)[,-1]
used <- merge(used, newframe, all.x = T)
#We have now calculated and named all of the variables for the model
surveydata<-used

```

```{r demographics, echo=FALSE, message=FALSE, warning=FALSE}
age_tab <- xtabs(~ cut(2020-Q37, breaks = c(0,24,34,44,54,64,74,120)) + RegionAbbr , fulldata)
names(attr(age_tab, "dimnames")) <- c("Age","State")
attr(age_tab,"dimnames")["Age"][[1]] <- c("Under 25", "25-35", "35-45", "45-55", "55-65", "65-75", "75+")
age_chi <- summary(age_tab)
age_final <- round(prop.table(addmargins(age_tab,2),2),3)*100

ethnicity <- xtabs( ~ factor(Q38, labels = c("Hispanic", "Non-Hispanic")) + RegionAbbr, fulldata)
names(attr(ethnicity, "dimnames")) <- c("Ethnicity","State")
ethnic_chi <- summary(ethnicity)
ethnic_final <- round(prop.table(addmargins(ethnicity,2),2),3)*100

politics <- xtabs( ~ polVec + RegionAbbr, used)
names(attr(politics, "dimnames")) <- c("Politics","State")
politics_chi <- summary(politics)
politics_final  <- round(prop.table(addmargins(politics,2),2),3)*100

sex <- xtabs( ~ factor(Q46, labels = c("Male","Female","Not Identified")) + RegionAbbr, fulldata)
names(attr(sex, "dimnames")) <- c("Gender","State")
sex_chi <- summary(sex)
sex_final  <- round(prop.table(addmargins(sex,2),2),3)*100

rurality <- xtabs( ~ factor(Urb_Rur, labels = c("Rural","Urban")) + RegionAbbr, fulldata)
names(attr(rurality, "dimnames")) <- c("Rurality","State")
rurality_chi <- summary(rurality)
rurality_final <- round(prop.table(addmargins(rurality,2),2),3)*100

religion <- xtabs( ~ factor(Q42, labels = c("Evangelical", "Catholic", "Mainline Protestant", "Latter-Day Saints", "Jewish", "Islam", "Buddhist", "Atheist", "Agnostic", "Other")) + RegionAbbr, fulldata)
names(attr(religion, "dimnames")) <- c("Religion","State")
religion_chi <- summary(religion)
religion_final <- round(prop.table(addmargins(religion,2),2),3)*100

race <- xtabs( ~ factor(Q39, labels = c("No Answer", "American Indian or Alaska Native", "Multi-race", "Multi-race", "Multi-race", "Asian or Asian-American", "Multi-race",
                                        "Multi-race", "Black or African-American", "Multi-race", "Native Hawaiian or Other Pacific Islander", "White", "Multi-race", "Other"))
               + RegionAbbr, fulldata)
names(attr(race, "dimnames")) <- c("Race","State")
race_chi <- summary(race)
race_final <- round(prop.table(addmargins(race,2),2),3)*100

income <- xtabs( ~ factor(Q45, labels = c("<$25,000", "$25,000-$49,999", "$50,000-$74,999", ">$74,999")) + RegionAbbr, fulldata)
names(attr(income, "dimnames")) <- c("Income Bracket","State")
income_chi <- summary(income)
income_final <- round(prop.table(addmargins(income,2),2),3)*100

relationship <- xtabs( ~ factor(Q44, labels = c("Single, never married", "Married or domestic partnership", "Widowed", "Divorced", "Separated")) + RegionAbbr, fulldata)
names(attr(relationship, "dimnames")) <- c("Relationship Status","State")
relationship_chi <- summary(relationship)
relationship_final <- round(prop.table(addmargins(relationship,2),2),3)*100

education <- xtabs( ~ factor(Q40, labels = c("Did not graduate HS", "HS diploma or equivalent", "Some college", "Associate's degree", "College graduate or higher")) + RegionAbbr, fulldata)
names(attr(education, "dimnames")) <- c("Educational Attainment","State")
education_chi <- summary(education)
education_final <- round(prop.table(addmargins(education,2),2),3)*100

demographics <- rbind(age_final, education_final,ethnic_final,sex_final,income_final,politics_final,race_final,relationship_final,religion_final,rurality_final)
demographics_group <- c("Age","Age","Age","Age","Age","Age","Age", "Ethnicity", "Ethnicity", "Race","Race","Race","Race","Race","Race","Race","Race","Religion","Religion","Religion","Religion","Religion","Religion","Religion","Religion","Religion","Religion","Gender","Gender","Gender","Politics","Politics","Politics","Politics","Politics","Politics","Rurality", "Rurality", "Educational Attainment", "Educational Attainment", "Educational Attainment", "Educational Attainment", "Educational Attainment", "Income Bracket", "Income Bracket", "Income Bracket", "Income Bracket", "Relationship Status", "Relationship Status", "Relationship Status", "Relationship Status", "Relationship Status")

```

\newpage 

## Table S3.1: Survey Demographics

```{r demographics_output, echo=FALSE, message=FALSE, warning=FALSE}

library(kableExtra)

age_text <- paste("Age: ", "chi sq. = ", round(age_chi$statistic, 2), ", df = ", age_chi$parameter, ", p-value = ", round(age_chi$p.value, 3), sep="")
educ_text <- paste("Educational Attainment: ", "chi sq. = ", round(education_chi$statistic, 2), ", df = ", education_chi$parameter, ", p-value = ", round(education_chi$p.value, 3), sep="")
ethnicity_text <- paste("Ethnicity: ", "chi sq. = ", round(ethnic_chi$statistic, 2), ", df = ", ethnic_chi$parameter, ", p-value = ", round(ethnic_chi$p.value,3), sep="")
race_text <- paste("Race: ", "chi sq. = ", round(race_chi$statistic, 2), ", df = ", race_chi$parameter, ", p-value = ", round(race_chi$p.value,3), sep="")
religion_text <- paste("Religion: ", "chi sq. = ", round(religion_chi$statistic, 2), ", df = ", religion_chi$parameter, ", p-value = ", round(religion_chi$p.value, 3), sep="")
gender_text <- paste("Gender: ", "chi sq. = ", round(sex_chi$statistic, 2), ", df = ", sex_chi$parameter, ", p-value = ", round(sex_chi$p.value, 3), sep="")
income_text <- paste("Income Bracket: ", "chi sq. = ", round(income_chi$statistic, 2), ", df = ", income_chi$parameter, ", p-value = ", round(income_chi$p.value, 3), sep="")
politics_text <- paste("Politics: ", "chi sq. = ", round(politics_chi$statistic, 2), ", df = ", politics_chi$parameter, ", p-value = ", round(politics_chi$p.value,3), sep="")
rurality_text <- paste("Rurality: ", "chi sq. = ", round(rurality_chi$statistic, 2), ", df = ", rurality_chi$parameter, ", p-value = ", round(rurality_chi$p.value,3), sep="")
relationship_text <- paste("Relationship Status: ", "chi sq. = ", round(relationship_chi$statistic, 2), ", df = ", relationship_chi$parameter, ", p-value = ", round(relationship_chi$p.value,3), sep="")

#plotting demographics table
kable(demographics, linesep = "", col.names = c("ID Pct", "TX Pct", "VT Pct", "Total Pct"), longtable = T) %>% 
  pack_rows(index = table(demographics_group)) %>% 
kable_styling(full_width = F, 
                position = "left", latex_options = "repeat_header")   %>% footnote(general = c(paste("Totals: ID: n = 526", "   TX: n = 143", "  VT: n = 365 ", sep=" "),
                                                                                               age_text, educ_text, ethnicity_text, gender_text, income_text, politics_text,
                                                                                               race_text, relationship_text, religion_text, rurality_text))

```

\newpage

## Table S3.2: RUCA Code Descriptions

```{r ruca_output, echo=FALSE, message=FALSE, warning=FALSE}
library(kableExtra)

RUCA_description <- read.csv("./data/RUCA_description.csv")
colnames(RUCA_description) <- c("Primary RUCA Codes", "Description")
kable(RUCA_description) %>% kable_styling(full_width = F, 
                position = "center", font_size = 8, latex_options = "hold_position")
```


\newpage

## Figure S3.1: Conceptual Models

![](./figures/figure_S1_survey_concept_models.png){#id .class width=100% height=100%}  

## Figure S3.2: Behavioral Intention Latent Variable Input

![](./figures/figure_S2_behavior_latent.png){#id .class width=100% height=100%}  

\newpage
## Figure S3.3: Trust, Health Risk Perception, and Economic Risk Perception Latent Variable Input

![](./figures/figure_S3_trust_risk_latent.png){#id .class width=100% height=100%}  

```{r DiseaseBurdenOutcome_1, echo=FALSE, message=FALSE, warning=FALSE, results=FALSE, include=FALSE}
trust_risk_burden_cases <- '
      # latent variable definitions
      trust =~ Q26_1+Q26_2+Q26_3 + Q26_5+Q26_6+Q26_7+Q26_8
      health_risk =~ Q2_1+Q2_2+Q2_3+Q3_1+Q3_5+Q3_6
      econ_risk =~ Q3_2+Q3_3+Q3_4+Q3_7
      #behavioral breakdown
      indoor_close =~ Q4_1 + Q5_1 + Q6_1 + Q7_1 + Q8_1
      indoor_group =~ Q4_2 + Q5_2 + Q6_2 + Q7_2 + Q8_2
      restaurant =~ Q4_3 + Q5_3 + Q6_3 + Q7_3 + Q8_3
      church =~ Q4_4 + Q5_4 + Q6_4 + Q7_4 + Q8_4
      shop =~ Q4_5 + Q5_5 + Q6_5 + Q7_5 + Q8_5
      appointments =~ Q4_6 + Q5_6 + Q6_6 + Q7_6 + Q8_6
      community =~ Q4_7 + Q5_7 + Q6_7 + Q7_7 + Q8_7
      mask =~  Q5_8 + Q6_8 + Q7_8 + Q8_8
      beh_int =~ indoor_close + indoor_group + restaurant + church + shop + appointments + community + mask 
      
      # regressions
     trust ~ h*ruralbool + m*femalebool + whitebool + age65plusbool + polVecLiberal + polVecConservative + polVecLibertarian + polVecNonpolitical + polVecOther
     health_risk ~ a*trust + q*ruralbool + n*femalebool + whitebool + l*age65plusbool + polVecLiberal + polVecConservative + polVecLibertarian + polVecNonpolitical + polVecOther
     econ_risk ~ c*trust +j*ruralbool + femalebool + whitebool + age65plusbool + polVecLiberal + polVecConservative + polVecLibertarian + polVecNonpolitical + polVecOther
     beh_int ~ e*econ_risk + f*health_risk + p*ruralbool + femalebool + whitebool + age65plusbool + polVecLiberal + polVecConservative + polVecLibertarian + polVecNonpolitical + polVecOther
     cumcasesper100 ~ g*beh_int + k*ruralbool 
     
     
      Trust := a*f*g #trust in governemnt effect on disease burden
      Health_risk := f*g #health risk effect on disease burden
      Rural_via_trust := h*a*f*g
'
 
caseburden <- sem(model=trust_risk_burden_cases,data=surveydata, std.ov=T)

```

## Table S3.3: Model 1A - Trust to Risk Perception on Disease Burden Summary Output Table


```{r, echo= FALSE, message = FALSE, warning = FALSE, error = FALSE, results = FALSE, include=FALSE}


fit <- summary(caseburden, fit.measures = TRUE, standardized = TRUE)

model_table <- fit$PE[c(57:106, 221:223),]
model_table <- as.data.frame(model_table)
model_table$est <- round(as.numeric(model_table$std.all), 3)
model_table$se <- round(as.numeric(model_table$se), 3)
model_table$z <- round(as.numeric(model_table$z), 3)
model_table$pvalue <- round(as.numeric(model_table$pvalue), 3)
model_table <- model_table[,c(1,3,11,7,8,9)]
colnames(model_table) <- c("Latent Variable", "Regression Variable", "std.est", "std.error", "z", "p-value")

model_table_original <- model_table

model_table$`Latent Variable`[2:8] <- " "
model_table$`Latent Variable`[10:17] <- " "
model_table$`Latent Variable`[19:27] <- " "
model_table$`Latent Variable`[29:37] <- " "
model_table$`Latent Variable`[39:48] <- " "
model_table$`Latent Variable`[50] <- " "

model_table$`Regression Variable`[51:53] <- " "


```


```{r, echo= FALSE, message = FALSE, warning = FALSE}

rownames(model_table) <- NULL
library(kableExtra)


kable(model_table, linesep = "")%>%
kable_styling(full_width = F,
                position = "left", font_size = 8) %>%footnote(general = paste("chisq = ", round(fit$FIT[3], 3),  "  aic = ", round(fit$FIT[10], 3), "  rmsea = ", round(fit$FIT[14], 3), sep=""))

# model_table_word <- read_docx(path = "./table_templates/COVID_survey_table_template.docx") %>% body_add_table(head(model_table, n = 33, style = "table_template")) %>% body_add_break()
#   
# caption <- block_caption(paste("chisq = ", round(fit$FIT[3], 3), "  cfi = ",  round(fit$FIT[9], 3), "  aic = ", round(fit$FIT[13], 3), "  rmsea = ", round(fit$FIT[17], 3), sep=""), style = "Normal")
# 
#   body_add_caption(model_table_word,  caption)
  
```
  

```{r, echo= FALSE, message = FALSE, warning = FALSE, include = FALSE}

# print(model_table_word, target = "./tables/Model3_table.docx")
```

## Figure S3.4: Model 1A - Trust to Risk Perception on Disease Burden Full Diagram


```{r, echo= FALSE, message = FALSE, warning = FALSE }

obj <- do.call(semPlotModel, list(caseburden)) # this is more general / not just for Mplus
idx <- apply(obj@Pars[c("lhs", "rhs")], 1, function(i) any(grepl("Q",  i)))
obj@Pars <- obj@Pars[!idx, ]

model_table_diagram <- model_table
model_table_diagram[is.na(model_table_diagram)] = 0
model_table_diagram$color <- ifelse(model_table_diagram$std.est > 0,                  # First test condition
 
       yes = ifelse(model_table_diagram$`p-value` > .05 & model_table_diagram$`p-value` < .1,    # Second test condition
                    yes =  "gray",
                    no = "blue"),
 
       no = ifelse(model_table_diagram$`p-value` > .05 & model_table_diagram$`p-value` < .1,    # Second test condition
                    yes = "gray",
                    no = "red"))

model_table_diagram$sig_binary <- ifelse(model_table_diagram$`p-value` > .1, 1,0)
model_table_diagram <- model_table_diagram[-c(51:53),]
obj@Pars <- obj@Pars[1:50,]

rownames(model_table_diagram) <- as.numeric(rownames(model_table_diagram))+56


len <- rownames(model_table_diagram[model_table_diagram$sig_binary == 0,])
#obj@Pars <- obj@Pars[rownames(obj@Pars) == rownames(model_table_diagram[model_table_diagram$sig_binary == 0,]), ]   

obj@Pars <- obj@Pars[rownames(obj@Pars) %in% len, ] 


model_table_diagram_refined <- model_table_diagram[model_table_diagram$sig_binary == 0,]


#--create custom layout
ly <- t(matrix(c(3,3,8,3,10,3,7,3,5,3,6,3,4,3,9,3,8,5,5,1,2,3,1,3,9,7,8,7,7,7,6,7,5,7,4,7,3,7,2,7,5,5),2))


semPlot::semPaths(obj, bifactor = "g", fade = FALSE, style = "lisrel", label.cex = .8, nCharNodes = 0,  what = "std", layout=ly, curvePivot = TRUE, edge.label.cex=1, intercepts = FALSE, sizeLat = 9, sizeLat2 =6, rotation = 3, residuals = FALSE, exoVar = FALSE, exoCov = FALSE, label.scale=FALSE, edgeLabels = FALSE, sizeMan = 8, sizeMan2 = 4, allVars = FALSE, mar = c(1.5,1.1,1.5,1.1), thresholds = FALSE, optimizeLatRes = TRUE, curve = c(0,0,0,0,0,0,0,0,0,0,0,0,0,0,0,-1,-1,1,1,-1,0,-1,1,1,-1,-1,0,0,0,0,0,0,0), edge.color = model_table_diagram_refined$color, nodeLabels = c("", "", "", "", "", "", "", "", "", "", "", "", "", "", "", "", "", "", "", "", "", "", "", "", "", "", "", "", "", "", "", "", "", "", "", "", "", "", "", "", "", "", "", "", "", "", "", "", "", "", "", "", "D", "C", "B", "A",     "NONPOL", "WHITE","RURAL", "AGE 65+","LIB", "LIBERTRN", "CONS", "FEMALE", "CASES",  "", "TRUST", "HEALTH\nRISK", "ECONOMIC\nRISK", "INDOOR\nCLOSE", "INDOOR\nGROUP", "RESTRNTS", "CHURCH", "SHOP", "APPTS", "COMMUNITY", "MASK",  "BEHAVIOR", "", "", "" ))

legend("bottomright",
       legend = c("negative coefficient", "positive coefficent", "marginally non-significant\ncoefficient (0.05 < p < 0.1)"),
       lty = c(5, 5, 5), col = c("red", "blue", "gray"), lwd = 5, cex = 1.1)


legend("bottomleft",
       legend = c("observed variable", "latent variable"),
        col = c("black"), pch = c(0,1), cex = 1.1)

#summary(separatedtypes, fit.measures = TRUE)

```


## Figure S3.5: Model 1A - Trust to Risk Perception on Disease Burden Simplified Diagram

```{r, echo= FALSE, message = FALSE, warning = FALSE }

obj <- do.call(semPlotModel, list(caseburden)) # this is more general / not just for Mplus
#idx <- apply(obj@Pars[c("lhs", "rhs")], 1, function(i) any(grepl(c("bool|Q|deaths|cases|mask|community|shopping|appointments|shop|restaurant|indoor_group|indoor_close|church"),  i)))
#obj@Pars <- rbind(obj@Pars[!idx, ], 
#obj@Pars <- obj@Pars[c(65,69,74:75,79,84,94:96, 100,105:106),]

model_table_simplified <- model_table_original[c("65","74","75","84","94", "95", "96", "105", "106"),]
model_table_simplified$color <- ifelse(model_table_simplified$std.est > 0,                  # First test condition
 
       yes = ifelse(model_table_simplified$`p-value` > .05,    # Second test condition
                    yes =  "gray",
                    no = "blue"),
 
       no = ifelse(model_table_simplified$`p-value` > .05,    # Second test condition
                    yes = "gray",
                    no = "red"))

model_table_simplified$sig_binary <- ifelse(model_table_simplified$`p-value` > .1, 1,0)
model_table_simplified_refined <- model_table_simplified[model_table_simplified$sig_binary == 0,]


obj@Pars <- obj@Pars[rownames(model_table_simplified_refined),]
ly <- t(matrix(c(4,3,4,2,1,2,2,1,2,3,3,2), 2))


semPlot::semPaths(obj, bifactor = "g", fade = FALSE, style = "lisrel", label.cex = .8, nCharNodes = 0,  what = "std", layout=ly, curvePivot = TRUE, edge.label.cex=1, intercepts = FALSE, sizeLat = 14, sizeLat2 =8, rotation = 3, residuals = FALSE, exoVar = FALSE, exoCov = FALSE, label.scale=FALSE, layoutSplit = TRUE, edgeLabels = TRUE, sizeMan = 10, sizeMan2 = 5, allVars = FALSE, mar = c(7,3,7,3), thresholds = FALSE, optimizeLatRes = TRUE, curve = c(0,-1,1,-1,-1,0), edge.color = model_table_simplified_refined$color, nodeLabels = c("","","","","","","","","","","","","","","","","","","","","","","","","","","","","","","","","","","","","","","","","","","","","","","","","","","","","","","","","RURALITY","CASES","","","","CASES","","","","","TRUST","HEALTH\nRISK\nPERCEPTION","ECONOMIC\nRISK\nPERCEPTION","g","h","i","","","","","","BEHAVIORAL\nINTENTION"))

#edge.color = c("darkgray", "blue", "blue", "red", "gray", "blue", "red", "red", "red", "red", "red", "red"), 

legend("bottomright",
       legend = c("negative coefficient", "positive coefficent", "marginally non-significant\ncoefficient (0.05 < p < 0.1)"),
       lty = c(5, 5, 5), col = c("red", "blue", "gray"), lwd = 5, cex = 1.1)


legend("bottomleft",
       legend = c("observed variable", "latent variable"),
        col = c("black"), pch = c(0,1), cex = 1.1)

#summary(separatedtypes, fit.measures = TRUE)

```


```{r DiseaseBurdenOutcome_3, echo=FALSE, message=FALSE, warning=FALSE, results=FALSE, include=FALSE}
risktotrust_burden_cases <- '
      # latent variable definitions
      trust =~ Q26_1+Q26_2+Q26_3 + Q26_5+Q26_6+Q26_7+Q26_8
      health_risk =~ Q2_1+Q2_2+Q2_3+Q3_1+Q3_5+Q3_6
      econ_risk =~ Q3_2+Q3_3+Q3_4+Q3_7
      #behavioral breakdown
      indoor_close =~ Q4_1 + Q5_1 + Q6_1 + Q7_1 + Q8_1
      indoor_group =~ Q4_2 + Q5_2 + Q6_2 + Q7_2 + Q8_2
      restaurant =~ Q4_3 + Q5_3 + Q6_3 + Q7_3 + Q8_3
      church =~ Q4_4 + Q5_4 + Q6_4 + Q7_4 + Q8_4
      shop =~ Q4_5 + Q5_5 + Q6_5 + Q7_5 + Q8_5
      appointments =~ Q4_6 + Q5_6 + Q6_6 + Q7_6 + Q8_6
      community =~ Q4_7 + Q5_7 + Q6_7 + Q7_7 + Q8_7
      mask =~  Q5_8 + Q6_8 + Q7_8 + Q8_8
      beh_int =~ indoor_close + indoor_group + restaurant + church + shop + appointments + community + mask 
      
      # regressions
     trust ~ ruralbool + femalebool + whitebool + age65plusbool + polVecLiberal + polVecConservative + polVecLibertarian + polVecNonpolitical + polVecOther + health_risk + econ_risk
     health_risk ~  ruralbool + femalebool + whitebool + age65plusbool + polVecLiberal + polVecConservative + polVecLibertarian + polVecNonpolitical + polVecOther
     econ_risk ~ ruralbool + femalebool + whitebool + age65plusbool + polVecLiberal + polVecConservative + polVecLibertarian + polVecNonpolitical + polVecOther
     beh_int ~  trust +ruralbool + femalebool + whitebool + age65plusbool + polVecLiberal + polVecConservative + polVecLibertarian + polVecNonpolitical + polVecOther 
     cumcasesper100 ~ beh_int + ruralbool 
     
'
 
caseburdenrtot <- sem(model=risktotrust_burden_cases,data=surveydata, std.ov=T)
```


## Table S3.4: Model 2A Risk Perception to Trust on Disease Burden Summary Table Output

```{r, echo= FALSE, message = FALSE, warning = FALSE, error = FALSE, results = FALSE, include=FALSE}

fit <- summary(caseburdenrtot, fit.measures = TRUE, standardized = TRUE)


model_table <- fit$PE[c(57:105),]
model_table <- as.data.frame(model_table)
model_table$est <- round(as.numeric(model_table$std.all), 3)
model_table$se <- round(as.numeric(model_table$se), 3)
model_table$z <- round(as.numeric(model_table$z), 3)
model_table$pvalue <- round(as.numeric(model_table$pvalue), 3)
model_table <- model_table[,c(1,3,10,6,7,8)]
colnames(model_table) <- c("Latent Variable", "Regression Variable", "std.est", "std.error", "z", "p-value")

model_table_original <- model_table

model_table$`Latent Variable`[2:8] <- " "
model_table$`Latent Variable`[10:19] <- " "
model_table$`Latent Variable`[21:28] <- " "
model_table$`Latent Variable`[30:37] <- " "
model_table$`Latent Variable`[39:47] <- " "
model_table$`Latent Variable`[49] <- " "


```


```{r, echo= FALSE, message = FALSE, warning = FALSE}

rownames(model_table) <- NULL
library(kableExtra)


kable(model_table, linesep = "")%>%
kable_styling(full_width = F,
                position = "left", font_size = 8) %>%footnote(general = paste("chisq = ", round(fit$FIT[3], 3),  "  aic = ", round(fit$FIT[10], 3), "  rmsea = ", round(fit$FIT[14], 3), sep=""))

# model_table_word <- read_docx(path = "./table_templates/COVID_survey_table_template.docx") %>% body_add_table(head(model_table, n = 33, style = "table_template")) %>% body_add_break()
#   
# caption <- block_caption(paste("chisq = ", round(fit$FIT[3], 3), "  cfi = ",  round(fit$FIT[9], 3), "  aic = ", round(fit$FIT[13], 3), "  rmsea = ", round(fit$FIT[17], 3), sep=""), style = "Normal")
# 
#   body_add_caption(model_table_word,  caption)
  
```
  

```{r, echo= FALSE, message = FALSE, warning = FALSE, include = FALSE}

# print(model_table_word, target = "./tables/Model3_table.docx")
```

## Figure S3.6: Model 2A - Risk Perception to Trust on Disease Burden Full Diagram


```{r, echo= FALSE, message = FALSE, warning = FALSE }


obj <- do.call(semPlotModel, list(caseburdenrtot)) # this is more general / not just for Mplus
idx <- apply(obj@Pars[c("lhs", "rhs")], 1, function(i) any(grepl("Q",  i)))
obj@Pars <- obj@Pars[!idx, ]

model_table_diagram <- model_table
model_table_diagram[is.na(model_table_diagram)] = 0
model_table_diagram$color <- ifelse(model_table_diagram$std.est > 0,                  # First test condition
 
       yes = ifelse(model_table_diagram$`p-value` > .05 & model_table_diagram$`p-value` < .1,    # Second test condition
                    yes =  "gray",
                    no = "blue"),
 
       no = ifelse(model_table_diagram$`p-value` > .05 & model_table_diagram$`p-value` < .1,    # Second test condition
                    yes = "gray",
                    no = "red"))

model_table_diagram$sig_binary <- ifelse(model_table_diagram$`p-value` > .1, 1,0)
#model_table_diagram <- model_table_diagram[-c(51:53),]
obj@Pars <- obj@Pars[1:49,]

rownames(model_table_diagram) <- as.numeric(rownames(model_table_diagram))+56


len <- rownames(model_table_diagram[model_table_diagram$sig_binary == 0,])
#obj@Pars <- obj@Pars[rownames(obj@Pars) == rownames(model_table_diagram[model_table_diagram$sig_binary == 0,]), ]   

obj@Pars <- obj@Pars[rownames(obj@Pars) %in% len, ] 


model_table_diagram_refined <- model_table_diagram[model_table_diagram$sig_binary == 0,]


#--create custom layout
ly <- t(matrix(c(9,3,3,3,6,3,10,3,8,3,7,3,4,3,5,3,7,5,1,3,6,1,4,1,9,7,8,7,7,7,6,7,5,7,4,7,3,7,2,7,5,5),2))


semPlot::semPaths(obj, bifactor = "g", fade = FALSE, style = "lisrel", label.cex = .8, nCharNodes = 0,  what = "std", layout=ly, curvePivot = TRUE, edge.label.cex=1, intercepts = FALSE, sizeLat = 9, sizeLat2 =6, rotation = 3, residuals = FALSE, exoVar = FALSE, exoCov = FALSE, label.scale=FALSE, edgeLabels = FALSE, sizeMan = 8, sizeMan2 = 4, allVars = FALSE, mar = c(1.5,1.1,1.5,1.1), thresholds = FALSE, optimizeLatRes = TRUE,  curve = c(0,0,0,0,0,0,0,0,-1,-1,-1,-1,1,1,0,0,0,0,0,0,0,0,0,0,0,0,0,0,0,0,0), edge.color = model_table_diagram_refined$color, nodeLabels = c("", "", "", "", "", "", "", "", "", "", "", "", "", "", "", "", "", "", "", "", "", "", "", "", "", "", "", "", "", "", "", "", "", "", "", "", "", "", "", "", "", "", "", "", "", "", "", "", "", "", "", "", "D", "C", "B", "A",    "FEMALE", "NONPOL", "LIBERTRN", "RURAL","WHITE", "AGE 65+", "CONS", "LIB", "CASES",  "", "TRUST", "HEALTH\nRISK", "ECONOMIC\nRISK", "INDOOR\nCLOSE", "INDOOR\nGROUP", "RESTRNTS", "CHURCH", "SHOP", "APPTS", "COMMUNITY", "MASK",  "BEHAVIOR", "E", "F", "G" ))

legend("bottomright",
       legend = c("negative coefficient", "positive coefficent", "marginally non-significant\ncoefficient (0.05 < p < 0.1)"),
       lty = c(5, 5, 5), col = c("red", "blue", "gray"), lwd = 5, cex = 1.1)


legend("bottomleft",
       legend = c("observed variable", "latent variable"),
        col = c("black"), pch = c(0,1), cex = 1.1)

#summary(separatedtypes, fit.measures = TRUE)


```


## Figure S3.7: Model 2A - Risk Perception to Trust on Disease Burden Simplified Diagram

```{r, echo= FALSE, message = FALSE, warning = FALSE }


obj <- do.call(semPlotModel, list(caseburdenrtot)) # this is more general / not just for Mplus

model_table_simplified <- model_table_original[c("65","74", "75", "76", "85","94", "95", "104","105"),]
model_table_simplified$color <- ifelse(model_table_simplified$std.est > 0,                  # First test condition
 
       yes = ifelse(model_table_simplified$`p-value` > .05,    # Second test condition
                    yes =  "gray",
                    no = "blue"),
 
       no = ifelse(model_table_simplified$`p-value` > .05,    # Second test condition
                    yes = "gray",
                    no = "red"))

model_table_simplified$sig_binary <- ifelse(model_table_simplified$`p-value` > .1, 1,0)
model_table_simplified_refined <- model_table_simplified[model_table_simplified$sig_binary == 0,]


obj@Pars <- obj@Pars[rownames(model_table_simplified_refined),]
ly <- t(matrix(c(4,3,4,2,2,2,1,2,1,3,3,2), 2))


semPlot::semPaths(obj, bifactor = "g", fade = FALSE, style = "lisrel", label.cex = .8, nCharNodes = 0,  what = "std", layout=ly, curvePivot = TRUE, edge.label.cex=1, intercepts = FALSE, sizeLat = 14, sizeLat2 =8, rotation = 3, residuals = FALSE, exoVar = FALSE, exoCov = FALSE, label.scale=FALSE, layoutSplit = TRUE, edgeLabels = TRUE, sizeMan = 10, sizeMan2 = 5, allVars = FALSE, mar = c(3,3,3,3), thresholds = FALSE, optimizeLatRes = TRUE, curve = c(0,0,0,0,0,0), edge.color = model_table_simplified_refined$color, nodeLabels = c("","","","","","","","","","","","","","","","","","","","","","","","","","","","","","","","","","","","","","","","","","","","","","","","","","","","","","","","","RURALITY","CASES","","","","","","","","","TRUST","HEALTH\nRISK\nPERCEPTION","ECONOMIC\nRISK\nPERCEPTION","","","","","","","","","BEHAVIORAL\nINTENTION"))

#edge.color = c("darkgray", "blue", "blue", "red", "gray", "blue", "red", "red", "red", "red", "red", "red"), 

legend("bottomright",
       legend = c("negative coefficient", "positive coefficent", "marginally non-significant\ncoefficient (0.05 < p < 0.1)"),
       lty = c(5, 5, 5), col = c("red", "blue", "gray"), lwd = 5, cex = 1.1)


legend("bottomleft",
       legend = c("observed variable", "latent variable"),
        col = c("black"), pch = c(0,1), cex = 1.1)

#summary(separatedtypes, fit.measures = TRUE)


```


```{r DiseaseBurdenOutcome_2, echo=FALSE, message=FALSE, warning=FALSE, results=FALSE, include=FALSE}
trustandrisk_burden_cases <- '
      # latent variable definitions
      trust =~ Q26_1+Q26_2+Q26_3 + Q26_5+Q26_6+Q26_7+Q26_8
      health_risk =~ Q2_1+Q2_2+Q2_3+Q3_1+Q3_5+Q3_6
      econ_risk =~ Q3_2+Q3_3+Q3_4+Q3_7
      #behavioral breakdown
      indoor_close =~ Q4_1 + Q5_1 + Q6_1 + Q7_1 + Q8_1
      indoor_group =~ Q4_2 + Q5_2 + Q6_2 + Q7_2 + Q8_2
      restaurant =~ Q4_3 + Q5_3 + Q6_3 + Q7_3 + Q8_3
      church =~ Q4_4 + Q5_4 + Q6_4 + Q7_4 + Q8_4
      shop =~ Q4_5 + Q5_5 + Q6_5 + Q7_5 + Q8_5
      appointments =~ Q4_6 + Q5_6 + Q6_6 + Q7_6 + Q8_6
      community =~ Q4_7 + Q5_7 + Q6_7 + Q7_7 + Q8_7
      mask =~  Q5_8 + Q6_8 + Q7_8 + Q8_8
      beh_int =~ indoor_close + indoor_group + restaurant + church + shop + appointments + community + mask 
      
      # regressions
     trust ~ ruralbool + femalebool + whitebool + age65plusbool + polVecLiberal + polVecConservative + polVecLibertarian + polVecNonpolitical + polVecOther
     health_risk ~  ruralbool + femalebool + whitebool + age65plusbool + polVecLiberal + polVecConservative + polVecLibertarian + polVecNonpolitical + polVecOther
     econ_risk ~ ruralbool + femalebool + whitebool + age65plusbool + polVecLiberal + polVecConservative + polVecLibertarian + polVecNonpolitical + polVecOther
     beh_int ~ econ_risk + health_risk + trust +ruralbool + femalebool + whitebool + age65plusbool + polVecLiberal + polVecConservative + polVecLibertarian + polVecNonpolitical + polVecOther 
     cumcasesper100 ~ beh_int + ruralbool 
     
'
 
caseburdentandr <- sem(model=trustandrisk_burden_cases,data=surveydata, std.ov=T)
```

## Table S3.5: Model 3A Trust and Risk Perception on Disease Burden Summary Table Output

```{r, echo= FALSE, message = FALSE, warning = FALSE, error = FALSE, results = FALSE, include=FALSE}


fit <- summary(caseburdentandr, fit.measures = TRUE, standardized = TRUE)


model_table <- fit$PE[c(57:105),]
model_table <- as.data.frame(model_table)
model_table$est <- round(as.numeric(model_table$std.all), 3)
model_table$se <- round(as.numeric(model_table$se), 3)
model_table$z <- round(as.numeric(model_table$z), 3)
model_table$pvalue <- round(as.numeric(model_table$pvalue), 3)
model_table <- model_table[,c(1,3,10,6,7,8)]
colnames(model_table) <- c("Latent Variable", "Regression Variable", "std.est", "std.error", "z", "p-value")

model_table_original <- model_table

model_table$`Latent Variable`[2:8] <- " "
model_table$`Latent Variable`[10:17] <- " "
model_table$`Latent Variable`[19:26] <- " "
model_table$`Latent Variable`[28:35] <- " "
model_table$`Latent Variable`[37:47] <- " "
model_table$`Latent Variable`[39] <- " "


```


```{r, echo= FALSE, message = FALSE, warning = FALSE}

rownames(model_table) <- NULL
library(kableExtra)


kable(model_table, linesep = "")%>%
kable_styling(full_width = F,
                position = "left", font_size = 8) %>%footnote(general = paste("chisq = ", round(fit$FIT[3], 3),  "  aic = ", round(fit$FIT[10], 3), "  rmsea = ", round(fit$FIT[14], 3), sep=""))

# model_table_word <- read_docx(path = "./table_templates/COVID_survey_table_template.docx") %>% body_add_table(head(model_table, n = 33, style = "table_template")) %>% body_add_break()
#   
# caption <- block_caption(paste("chisq = ", round(fit$FIT[3], 3), "  cfi = ",  round(fit$FIT[9], 3), "  aic = ", round(fit$FIT[13], 3), "  rmsea = ", round(fit$FIT[17], 3), sep=""), style = "Normal")
# 
#   body_add_caption(model_table_word,  caption)
  
```
  

```{r, echo= FALSE, message = FALSE, warning = FALSE, include = FALSE}

# print(model_table_word, target = "./tables/Model3_table.docx")
```

## Figure S3.8: Model 3A - Trust and Risk Perception on Disease Burden Full Diagram


```{r, echo= FALSE, message = FALSE, warning = FALSE }


obj <- do.call(semPlotModel, list(caseburdentandr)) # this is more general / not just for Mplus
idx <- apply(obj@Pars[c("lhs", "rhs")], 1, function(i) any(grepl("Q",  i)))
obj@Pars <- obj@Pars[!idx, ]

model_table_diagram <- model_table
model_table_diagram[is.na(model_table_diagram)] = 0
model_table_diagram$color <- ifelse(model_table_diagram$std.est > 0,                  # First test condition
 
       yes = ifelse(model_table_diagram$`p-value` > .05 & model_table_diagram$`p-value` < .1,    # Second test condition
                    yes =  "gray",
                    no = "blue"),
 
       no = ifelse(model_table_diagram$`p-value` > .05 & model_table_diagram$`p-value` < .1,    # Second test condition
                    yes = "gray",
                    no = "red"))

model_table_diagram$sig_binary <- ifelse(model_table_diagram$`p-value` > .099, 1,0)
#model_table_diagram <- model_table_diagram[-c(51:53),]
obj@Pars <- obj@Pars[1:49,]


rownames(model_table_diagram) <- as.numeric(rownames(model_table_diagram))+56

len <- rownames(model_table_diagram[model_table_diagram$sig_binary == 0,])
#obj@Pars <- obj@Pars[rownames(obj@Pars) == rownames(model_table_diagram[model_table_diagram$sig_binary == 0,]), ]   

obj@Pars <- obj@Pars[rownames(obj@Pars) %in% len, ] 


model_table_diagram_refined <- model_table_diagram[model_table_diagram$sig_binary == 0,]


#--create custom layout
ly <- t(matrix(c(9,3,4,3,7,3,8,3,11,3,6,3,5,3,10,3,9,5,1,3,3,3,2,3,10,7,9,7,8,7,7,7,6,7,5,7,4,7,3,7,6,5),2))


semPlot::semPaths(obj, bifactor = "g", fade = FALSE, style = "lisrel", label.cex = .8, nCharNodes = 0,  what = "std", layout=ly, curvePivot = TRUE, edge.label.cex=1, intercepts = FALSE, sizeLat = 8, sizeLat2 =6, rotation = 3, residuals = FALSE, exoVar = FALSE, exoCov = FALSE, label.scale=FALSE, edgeLabels = FALSE, sizeMan = 7, sizeMan2 = 4, allVars = FALSE, mar = c(7,.95,7,.95), thresholds = FALSE, optimizeLatRes = TRUE,  curve = c(0,0,0,0,0,0,0,0,-1,-1,-1,-1,-1,-1,.35,-1,-1,1,.4,-1,-1,-1,0,0,0,0,0,0,0), edge.color = model_table_diagram_refined$color, nodeLabels = c("", "", "", "", "", "", "", "", "", "", "", "", "", "", "", "", "", "", "", "", "", "", "", "", "", "", "", "", "", "", "", "", "", "", "", "", "", "", "", "", "", "", "", "", "", "", "", "", "", "", "", "", "D", "C", "B", "A", "WHITE","NONPOL", "LIBERTRN", "AGE 65+","RURAL", "LIB", "CONS", "FEMALE", "CASES",  "", "TRUST", "HEALTH\nRISK", "ECONOMIC\nRISK", "INDOOR\nCLOSE", "INDOOR\nGROUP", "RESTRNTS", "CHURCH", "SHOP", "APPTS", "COMMUNITY", "MASK",  "BEHAVIOR", "", "", "" ))


legend("bottomright",
       legend = c("negative coefficient", "positive coefficent", "marginally non-significant\ncoefficient (0.05 < p < 0.1)"),
       lty = c(5, 5, 5), col = c("red", "blue", "gray"), lwd = 5, cex = 1.1)


legend("bottomleft",
       legend = c("observed variable", "latent variable"),
        col = c("black"), pch = c(0,1), cex = 1.1)

#summary(separatedtypes, fit.measures = TRUE)


```


## Figure S3.9: Model 3A - Trust and Risk Perception on Disease Burden Simplified Diagram

```{r, echo= FALSE, message = FALSE, warning = FALSE }


obj <- do.call(semPlotModel, list(caseburdentandr)) # this is more general / not just for Mplus
#idx <- apply(obj@Pars[c("lhs", "rhs")], 1, function(i) any(grepl(c("bool|Q|deaths|cases|mask|community|shopping|appointments|shop|restaurant|indoor_group|indoor_close|church"),  i)))
#obj@Pars <- rbind(obj@Pars[!idx, ], 
#obj@Pars <- obj@Pars[c(65,69,74:75,79,84,94:96, 100,105:106),]

model_table_simplified <- model_table_original[c("65", "74","83","92", "93", "94", "95", "104","105"),]
model_table_simplified$color <- ifelse(model_table_simplified$std.est > 0,                  # First test condition
 
       yes = ifelse(model_table_simplified$`p-value` > .05,    # Second test condition
                    yes =  "gray",
                    no = "blue"),
 
       no = ifelse(model_table_simplified$`p-value` > .05,    # Second test condition
                    yes = "gray",
                    no = "red"))

model_table_simplified$sig_binary <- ifelse(model_table_simplified$`p-value` > .099, 1,0)
model_table_simplified_refined <- model_table_simplified[model_table_simplified$sig_binary == 0,]


obj@Pars <- obj@Pars[rownames(model_table_simplified_refined),]
ly <- t(matrix(c(4,3,4,2,2,1,1,2,2,3,3,2), 2))


semPlot::semPaths(obj, bifactor = "g", fade = FALSE, style = "lisrel", label.cex = .8, nCharNodes = 0,  what = "std", layout=ly, curvePivot = TRUE, edge.label.cex=1, intercepts = FALSE, sizeLat = 14, sizeLat2 =8, rotation = 3, residuals = FALSE, exoVar = FALSE, exoCov = FALSE, label.scale=FALSE, layoutSplit = TRUE, edgeLabels = TRUE, sizeMan = 10, sizeMan2 = 5, allVars = FALSE, mar = c(7,3,7,3), thresholds = FALSE, optimizeLatRes = TRUE, curve = c(-1,0,0,0,0), edge.color = model_table_simplified_refined$color, nodeLabels = c("","","","","","","","","","","","","","","","","","","","","","","","","","","","","","","","","","","","","","","","","","","","","","","","","","","","","","","","","RURALITY","CASES","","","","","","","","","TRUST","HEALTH\nRISK\nPERCEPTION","ECONOMIC\nRISK\nPERCEPTION","","","","","","","","","BEHAVIORAL\nINTENTION"))

#edge.color = c("darkgray", "blue", "blue", "red", "gray", "blue", "red", "red", "red", "red", "red", "red"), 

legend("bottomright",
       legend = c("negative coefficient", "positive coefficent", "marginally non-significant\ncoefficient (0.05 < p < 0.1)"),
       lty = c(5, 5, 5), col = c("red", "blue", "gray"), lwd = 5, cex = 1.1)


legend("bottomleft",
       legend = c("observed variable", "latent variable"),
        col = c("black"), pch = c(0,1), cex = 1.1)

#summary(separatedtypes, fit.measures = TRUE)


```


```{r DiseasePrevalence_1, echo=FALSE, message=FALSE, warning=FALSE, results=FALSE}
prev_trust_risk_total <- '
      # latent variable definitions
      trust =~ Q26_1+Q26_2+Q26_3 + Q26_5+Q26_6+Q26_7+Q26_8
      health_risk =~ Q2_1+Q2_2+Q2_3+Q3_1+Q3_5+Q3_6
      econ_risk =~ Q3_2+Q3_3+Q3_4+Q3_7
      #behavioral breakdown
      indoor_close =~ Q4_1 + Q5_1 + Q6_1 + Q7_1 + Q8_1
      indoor_group =~ Q4_2 + Q5_2 + Q6_2 + Q7_2 + Q8_2
      restaurant =~ Q4_3 + Q5_3 + Q6_3 + Q7_3 + Q8_3
      church =~ Q4_4 + Q5_4 + Q6_4 + Q7_4 + Q8_4
      shop =~ Q4_5 + Q5_5 + Q6_5 + Q7_5 + Q8_5
      appointments =~ Q4_6 + Q5_6 + Q6_6 + Q7_6 + Q8_6
      community =~ Q4_7 + Q5_7 + Q6_7 + Q7_7 + Q8_7
      mask =~  Q5_8 + Q6_8 + Q7_8 + Q8_8
      beh_int =~ indoor_close + indoor_group + restaurant + church + shop + appointments + community + mask 
      
      # regressions
     trust ~ h*ruralbool + femalebool + whitebool + age65plusbool + polVecLiberal + polVecConservative + polVecLibertarian + polVecNonpolitical + polVecOther
     health_risk ~ a*trust + ruralbool + femalebool + whitebool + age65plusbool + polVecLiberal + polVecConservative + polVecLibertarian + polVecNonpolitical + polVecOther
     econ_risk ~ c*trust +j*ruralbool + femalebool + whitebool + age65plusbool + polVecLiberal + polVecConservative + polVecLibertarian + polVecNonpolitical + polVecOther
     beh_int ~ e*econ_risk + f*health_risk + ruralbool + femalebool + whitebool + age65plusbool + polVecLiberal + polVecConservative + polVecLibertarian + polVecNonpolitical + polVecOther + casetodateper100 
     
    #  T_total := a*f*g + c*e*g #trust in governemnt effect on disease burden
    #  Rh_total := f*g #health risk effect on disease burden
    #  Re_total := e*g #economic risk effect on disease burden
    #  UR_trust := h*T_total #impact of rural on burden via trust in science
    #  UR_econ := j*Re_total #impact of rural on burden via economic risk
    #  UR_total := UR_trust + UR_econ + k #net impact of rural on burden
'
 
prevtotalmodel <- sem(model=prev_trust_risk_total,data=surveydata, std.ov=T)
```

## Table S3.6: Model 1B - Trust to Risk Perception on Behavioral Intention Summary Table Output

```{r, echo= FALSE, message = FALSE, warning = FALSE, error = FALSE, results = FALSE, include=FALSE}

fit <- summary(prevtotalmodel, fit.measures = TRUE, standardized = TRUE)

model_table <- fit$PE[c(57:105),]
model_table <- as.data.frame(model_table)
model_table$est <- round(as.numeric(model_table$std.all), 3)
model_table$se <- round(as.numeric(model_table$se), 3)
model_table$z <- round(as.numeric(model_table$z), 3)
model_table$pvalue <- round(as.numeric(model_table$pvalue), 3)
model_table <- model_table[,c(1,3,11,7,8,9)]
colnames(model_table) <- c("Latent Variable", "Regression Variable", "std.est", "std.error", "z", "p-value")

model_table_original <- model_table

model_table$`Latent Variable`[2:8] <- " "
model_table$`Latent Variable`[10:17] <- " "
model_table$`Latent Variable`[19:27] <- " "
model_table$`Latent Variable`[29:37] <- " "
model_table$`Latent Variable`[39:49] <- " "


```


```{r, echo= FALSE, message = FALSE, warning = FALSE}

rownames(model_table) <- NULL
library(kableExtra)


kable(model_table, linesep = "")%>%
kable_styling(full_width = F,
                position = "left", font_size = 8) %>%footnote(general = paste("chisq = ", round(fit$FIT[3], 3),  "  aic = ", round(fit$FIT[10], 3), "  rmsea = ", round(fit$FIT[14], 3), sep=""))

# model_table_word <- read_docx(path = "./table_templates/COVID_survey_table_template.docx") %>% body_add_table(head(model_table, n = 33, style = "table_template")) %>% body_add_break()
#   
# caption <- block_caption(paste("chisq = ", round(fit$FIT[3], 3), "  cfi = ",  round(fit$FIT[9], 3), "  aic = ", round(fit$FIT[13], 3), "  rmsea = ", round(fit$FIT[17], 3), sep=""), style = "Normal")
# 
#   body_add_caption(model_table_word,  caption)
  
```
  


```{r, echo= FALSE, message = FALSE, warning = FALSE, include = FALSE}

# print(model_table_word, target = "./tables/Model3_table.docx")
```

## Figure S3.10: Model 1B - Trust to Risk Perception on Behavioral Intention Full Diagram


```{r, echo= FALSE, message = FALSE, warning = FALSE }


obj <- do.call(semPlotModel, list(prevtotalmodel)) # this is more general / not just for Mplus
idx <- apply(obj@Pars[c("lhs", "rhs")], 1, function(i) any(grepl("Q",  i)))
obj@Pars <- obj@Pars[!idx, ]

model_table_diagram <- model_table
model_table_diagram[is.na(model_table_diagram)] = 0
model_table_diagram$color <- ifelse(model_table_diagram$std.est > 0,                  # First test condition
 
       yes = ifelse(model_table_diagram$`p-value` > .05 & model_table_diagram$`p-value` < .1,    # Second test condition
                    yes =  "gray",
                    no = "blue"),
 
       no = ifelse(model_table_diagram$`p-value` > .05 & model_table_diagram$`p-value` < .1,    # Second test condition
                    yes = "gray",
                    no = "red"))

model_table_diagram$sig_binary <- ifelse(model_table_diagram$`p-value` > .1, 1,0)
#model_table_diagram <- model_table_diagram[-c(51:53),]
obj@Pars <- obj@Pars[1:49,]

rownames(model_table_diagram) <- as.numeric(rownames(model_table_diagram))+56


len <- rownames(model_table_diagram[model_table_diagram$sig_binary == 0,])
#obj@Pars <- obj@Pars[rownames(obj@Pars) == rownames(model_table_diagram[model_table_diagram$sig_binary == 0,]), ]   

obj@Pars <- obj@Pars[rownames(obj@Pars) %in% len, ] 


model_table_diagram_refined <- model_table_diagram[model_table_diagram$sig_binary == 0,]


#--create custom layout
ly <- t(matrix(c(3,3,10,3,8,3,7,3,5,3,6,3,4,3,9,3,7,5,5,1,2,3,1,3,9,7,8,7,7,7,6,7,5,7,4,7,3,7,2,7,5,5),2))


semPlot::semPaths(obj, bifactor = "g", fade = FALSE, style = "lisrel", label.cex = .8, nCharNodes = 0,  what = "std", layout=ly, curvePivot = TRUE, edge.label.cex=1, intercepts = FALSE, sizeLat = 9, sizeLat2 =6, rotation = 3, residuals = FALSE, exoVar = FALSE, exoCov = FALSE, label.scale=FALSE, edgeLabels = FALSE, sizeMan = 8, sizeMan2 = 4, allVars = FALSE, mar = c(1.5,1.1,1.5,1.1), thresholds = FALSE, optimizeLatRes = TRUE,  curve = c(0,0,0,0,0,0,0,0,0,0,0,0,0,0,0,-1,-1,1,1,-1,0,-1,1,1,1,-1,0,0,0,0,0,0,0), edge.color = model_table_diagram_refined$color, nodeLabels = c("", "", "", "", "", "", "", "", "", "", "", "", "", "", "", "", "", "", "", "", "", "", "", "", "", "", "", "", "", "", "", "", "", "", "", "", "", "", "", "", "", "", "", "", "", "", "", "", "", "", "", "", "D", "C", "B", "A",     "NONPOL", "RURAL","WHITE", "AGE 65+","LIB", "LIBERTRN", "CONS", "FEMALE", "",  "CASES", "TRUST", "HEALTH\nRISK", "ECONOMIC\nRISK", "INDOOR\nCLOSE", "INDOOR\nGROUP", "RESTRNTS", "CHURCH", "SHOPPING", "APPTS", "COMMUNITY", "MASK",  "BEHAVIOR", "E", "F", "G" ))

legend("bottomright",
       legend = c("negative coefficient", "positive coefficent", "marginally non-significant\ncoefficient (0.05 < p < 0.1)"),
       lty = c(5, 5, 5), col = c("red", "blue", "gray"), lwd = 5, cex = 1.1)


legend("bottomleft",
       legend = c("observed variable", "latent variable"),
        col = c("black"), pch = c(0,1), cex = 1.1)

#summary(separatedtypes, fit.measures = TRUE)


```


## Figure S3.11: Model 1B - Trust to Risk Perception on Behavioral Intention Simplified Diagram

```{r, echo= FALSE, message = FALSE, warning = FALSE }

obj <- do.call(semPlotModel, list(prevtotalmodel)) # this is more general / not just for Mplus
#idx <- apply(obj@Pars[c("lhs", "rhs")], 1, function(i) any(grepl(c("bool|Q|deaths|cases|mask|community|shopping|appointments|shop|restaurant|indoor_group|indoor_close|church"),  i)))
#obj@Pars <- rbind(obj@Pars[!idx, ], 


model_table_simplified <- model_table_original[c("65","74","75","84","85", "94", "95", "105"),]
model_table_simplified$color <- ifelse(model_table_simplified$std.est > 0,                  # First test condition
 
       yes = ifelse(model_table_simplified$`p-value` > .05,    # Second test condition
                    yes =  "gray",
                    no = "blue"),
 
       no = ifelse(model_table_simplified$`p-value` > .05,    # Second test condition
                    yes = "gray",
                    no = "red"))

model_table_simplified$sig_binary <- ifelse(model_table_simplified$`p-value` > .1, 1,0)
model_table_simplified_refined <- model_table_simplified[model_table_simplified$sig_binary == 0,]


obj@Pars <- obj@Pars[rownames(model_table_simplified_refined),]
ly <- t(matrix(c(4,3,4,2,1,2,2,1,2,3,3,2), 2))


# 
# obj@Pars <- obj@Pars[c(65,70,76,83),]
# 
# ly <- t(matrix(c(4,3,1,2,2,1,2,3,3,2), 2))

semPlot::semPaths(obj, bifactor = "g", fade = FALSE, style = "lisrel", label.cex = .8, nCharNodes = 0,  what = "std", layout=ly, curvePivot = TRUE, edge.label.cex=1, intercepts = FALSE, sizeLat = 14, sizeLat2 =8, rotation = 3, residuals = FALSE, exoVar = FALSE, exoCov = FALSE, label.scale=FALSE, layoutSplit = TRUE, edgeLabels = TRUE, sizeMan = 10, sizeMan2 = 5, allVars = FALSE, mar = c(7,3,7,3), thresholds = FALSE, optimizeLatRes = TRUE, curve = c(0,-1,1,-1,1,-1,0), edge.color = model_table_simplified_refined$color, nodeLabels = c("","","","","","","","","","","","","","","","","","","","","","","","","","","","","","","","","","","","","","","","","","","","","","","","","","","","","","","","","RURALITY","","","","LIBERAL","","","","","CASES","TRUST","HEALTH\nRISK\nPERCEPTION","ECONOMIC\nRISK\nPERCEPTION","","","","","", "", "", "CASES", "BEHAVIORAL\nINTENTION", "", "", ""))

legend("bottomright",
       legend = c("negative coefficient", "positive coefficent", "marginally non-significant\ncoefficient (0.05 < p < 0.1)"),
       lty = c(5, 5, 5), col = c("red", "blue", "gray"), lwd = 5, cex = 1.1)


legend("bottomleft",
       legend = c("observed variable", "latent variable"),
        col = c("black"), pch = c(0,1), cex = 1.1)

#summary(separatedtypes, fit.measures = TRUE)

```


```{r DiseasePrevalence_3, echo=FALSE, message=FALSE, warning=FALSE, results=FALSE, include=FALSE}
prev_risk_trust_total <- '
      # latent variable definitions
      trust =~ Q26_1+Q26_2+Q26_3 + Q26_5+Q26_6+Q26_7+Q26_8
      health_risk =~ Q2_1+Q2_2+Q2_3+Q3_1+Q3_5+Q3_6
      econ_risk =~ Q3_2+Q3_3+Q3_4+Q3_7
      #behavioral breakdown
      indoor_close =~ Q4_1 + Q5_1 + Q6_1 + Q7_1 + Q8_1
      indoor_group =~ Q4_2 + Q5_2 + Q6_2 + Q7_2 + Q8_2
      restaurant =~ Q4_3 + Q5_3 + Q6_3 + Q7_3 + Q8_3
      church =~ Q4_4 + Q5_4 + Q6_4 + Q7_4 + Q8_4
      shop =~ Q4_5 + Q5_5 + Q6_5 + Q7_5 + Q8_5
      appointments =~ Q4_6 + Q5_6 + Q6_6 + Q7_6 + Q8_6
      community =~ Q4_7 + Q5_7 + Q6_7 + Q7_7 + Q8_7
      mask =~  Q5_8 + Q6_8 + Q7_8 + Q8_8
      beh_int =~ indoor_close + indoor_group + restaurant + church + shop + appointments + community + mask 
      
      # regressions
     trust ~ h*ruralbool + femalebool + whitebool + age65plusbool + polVecLiberal + polVecConservative + polVecLibertarian + polVecNonpolitical + polVecOther + health_risk + econ_risk
     health_risk ~ ruralbool + femalebool + whitebool + age65plusbool + polVecLiberal + polVecConservative + polVecLibertarian + polVecNonpolitical + polVecOther
     econ_risk ~ j*ruralbool + femalebool + whitebool + age65plusbool + polVecLiberal + polVecConservative + polVecLibertarian + polVecNonpolitical + polVecOther
     
      beh_int ~  trust +ruralbool + femalebool + whitebool + age65plusbool + polVecLiberal + polVecConservative + polVecLibertarian + polVecNonpolitical + polVecOther  + casetodateper100 

     # beh_int ~ e*econ_risk + f*health_risk + ruralbool + femalebool + whitebool + age65plusbool + polVecLiberal + polVecConservative + polVecLibertarian + polVecNonpolitical + polVecOther + casetodateper100 
     
    #  T_total := a*f*g + c*e*g #trust in governemnt effect on disease burden
    #  Rh_total := f*g #health risk effect on disease burden
    #  Re_total := e*g #economic risk effect on disease burden
    #  UR_trust := h*T_total #impact of rural on burden via trust in science
    #  UR_econ := j*Re_total #impact of rural on burden via economic risk
    #  UR_total := UR_trust + UR_econ + k #net impact of rural on burden
'
 
prevtotalrtmodel <- sem(model=prev_risk_trust_total,data=surveydata, std.ov=T)
```


## Table S3.7: Model 2B - Risk Perception to Trust on Behavioral Intention Summary Table Ouput

```{r, echo= FALSE, message = FALSE, warning = FALSE, error = FALSE, results = FALSE, include=FALSE}


fit <- summary(prevtotalrtmodel, fit.measures = TRUE, standardized = TRUE)


model_table <- fit$PE[c(57:104),]
model_table <- as.data.frame(model_table)
model_table$est <- round(as.numeric(model_table$std.all), 3)
model_table$se <- round(as.numeric(model_table$se), 3)
model_table$z <- round(as.numeric(model_table$z), 3)
model_table$pvalue <- round(as.numeric(model_table$pvalue), 3)
model_table <- model_table[,c(1,3,11,7,8,9)]
colnames(model_table) <- c("Latent Variable", "Regression Variable", "std.est", "std.error", "z", "p-value")

model_table_original <- model_table

model_table$`Latent Variable`[2:8] <- " "
model_table$`Latent Variable`[10:19] <- " "
model_table$`Latent Variable`[21:28] <- " "
model_table$`Latent Variable`[30:37] <- " "
model_table$`Latent Variable`[39:48] <- " "
#model_table$`Latent Variable`[49] <- " "


```


```{r, echo= FALSE, message = FALSE, warning = FALSE}

rownames(model_table) <- NULL
library(kableExtra)


kable(model_table, linesep = "")%>%
kable_styling(full_width = F,
                position = "left", font_size = 8) %>%footnote(general = paste("chisq = ", round(fit$FIT[3], 3),  "  aic = ", round(fit$FIT[10], 3), "  rmsea = ", round(fit$FIT[14], 3), sep=""))

# model_table_word <- read_docx(path = "./table_templates/COVID_survey_table_template.docx") %>% body_add_table(head(model_table, n = 33, style = "table_template")) %>% body_add_break()
#   
# caption <- block_caption(paste("chisq = ", round(fit$FIT[3], 3), "  cfi = ",  round(fit$FIT[9], 3), "  aic = ", round(fit$FIT[13], 3), "  rmsea = ", round(fit$FIT[17], 3), sep=""), style = "Normal")
# 
#   body_add_caption(model_table_word,  caption)
  
```
  

```{r, echo= FALSE, message = FALSE, warning = FALSE, include = FALSE}

# print(model_table_word, target = "./tables/Model3_table.docx")
```

## Figure S3.14: Model 2B - Risk Perception to Trust on Behavioral Intention Full Diagram


```{r, echo= FALSE, message = FALSE, warning = FALSE }


obj <- do.call(semPlotModel, list(prevtotalrtmodel)) # this is more general / not just for Mplus
idx <- apply(obj@Pars[c("lhs", "rhs")], 1, function(i) any(grepl("Q",  i)))
obj@Pars <- obj@Pars[!idx, ]

model_table_diagram <- model_table
model_table_diagram[is.na(model_table_diagram)] = 0
model_table_diagram$color <- ifelse(model_table_diagram$std.est > 0,                  # First test condition
 
       yes = ifelse(model_table_diagram$`p-value` > .05 & model_table_diagram$`p-value` < .1,    # Second test condition
                    yes =  "gray",
                    no = "blue"),
 
       no = ifelse(model_table_diagram$`p-value` > .05 & model_table_diagram$`p-value` < .1,    # Second test condition
                    yes = "gray",
                    no = "red"))

model_table_diagram$sig_binary <- ifelse(model_table_diagram$`p-value` > .1, 1,0)
#model_table_diagram <- model_table_diagram[-c(51:53),]
obj@Pars <- obj@Pars[1:49,]

rownames(model_table_diagram) <- as.numeric(rownames(model_table_diagram))+56


len <- rownames(model_table_diagram[model_table_diagram$sig_binary == 0,])
#obj@Pars <- obj@Pars[rownames(obj@Pars) == rownames(model_table_diagram[model_table_diagram$sig_binary == 0,]), ]   

obj@Pars <- obj@Pars[rownames(obj@Pars) %in% len, ] 


model_table_diagram_refined <- model_table_diagram[model_table_diagram$sig_binary == 0,]


#--create custom layout
ly <- t(matrix(c(9,3,3,3,10,3,7,3,6,3,8,3,4,3,5,3,1,3,6,1,4,1,9,7,8,7,7,7,6,7,5,7,4,7,3,7,2,7,5,5),2))


semPlot::semPaths(obj, bifactor = "g", fade = FALSE, style = "lisrel", label.cex = .8, nCharNodes = 0,  what = "std", layout=ly, curvePivot = TRUE, edge.label.cex=1, intercepts = FALSE, sizeLat = 9, sizeLat2 =6, rotation = 3, residuals = FALSE, exoVar = FALSE, exoCov = FALSE, label.scale=FALSE, edgeLabels = FALSE, sizeMan = 8, sizeMan2 = 4, allVars = FALSE, mar = c(1.5,1.1,1.5,1.1), thresholds = FALSE, optimizeLatRes = TRUE,  curve = c(0,0,0,0,0,0,0,0,-1,-1,-1,1,1,0,0,0,0,0,0,0,0,0,0,0,0,0,0,0,0,0), edge.color = model_table_diagram_refined$color, nodeLabels = c("", "", "", "", "", "", "", "", "", "", "", "", "", "", "", "", "", "", "", "", "", "", "", "", "", "", "", "", "", "", "", "", "", "", "", "", "", "", "", "", "", "", "", "", "", "", "", "", "", "", "", "", "D", "C", "B", "A",    "FEMALE", "NONPOL", "RURAL", "AGE 65+","LIBERTRN", "WHITE", "CONS", "LIB", "RURAL",  "", "TRUST", "HEALTH\nRISK", "ECONOMIC\nRISK", "INDOOR\nCLOSE", "INDOOR\nGROUP", "RESTRNTS", "CHURCH", "SHOPPING", "APPTS", "COMMUNITY", "MASK",  "BEHAVIOR", "E", "F", "G" ))

legend("bottomright",
       legend = c("negative coefficient", "positive coefficent", "marginally non-significant\ncoefficient (0.05 < p < 0.1)"),
       lty = c(5, 5, 5), col = c("red", "blue", "gray"), lwd = 5, cex = 1.1)


legend("bottomleft",
       legend = c("observed variable", "latent variable"),
        col = c("black"), pch = c(0,1), cex = 1.1)


```


## Figure S3.15: Model 2B - Risk Perception to Trust on Behavioral Intention Simplified Diagram

```{r, echo= FALSE, message = FALSE, warning = FALSE }

obj <- do.call(semPlotModel, list(prevtotalrtmodel)) # this is more general / not just for Mplus
# idx <- apply(obj@Pars[c("lhs", "rhs")], 1, function(i) any(grepl("Q",  i)))
# obj@Pars <- obj@Pars[!idx, ]


model_table_simplified <- model_table_original[c("65", "74", "75", "76", "85", "94", "95",  "104"),]
model_table_simplified$color <- ifelse(model_table_simplified$std.est > 0,                  # First test condition
 
       yes = ifelse(model_table_simplified$`p-value` > .05,    # Second test condition
                    yes =  "gray",
                    no = "blue"),
 
       no = ifelse(model_table_simplified$`p-value` > .05,    # Second test condition
                    yes = "gray",
                    no = "red"))

model_table_simplified$sig_binary <- ifelse(model_table_simplified$`p-value` > .1, 1,0)
model_table_simplified_refined <- model_table_simplified[model_table_simplified$sig_binary == 0,]


obj@Pars <- obj@Pars[rownames(model_table_simplified_refined),]

ly <- t(matrix(c(4,3,2,2,1,3,1,1,3,2), 2))


semPlot::semPaths(obj, bifactor = "g", fade = FALSE, style = "lisrel", label.cex = .8, nCharNodes = 0,  what = "std", layout=ly, curvePivot = TRUE, edge.label.cex=1, intercepts = FALSE, sizeLat = 14, sizeLat2 =8, rotation = 3, residuals = FALSE, exoVar = FALSE, exoCov = FALSE, label.scale=FALSE, layoutSplit = TRUE, edgeLabels = TRUE, sizeMan = 10, sizeMan2 = 5, allVars = FALSE, mar = c(7,3,7,3), thresholds = FALSE, optimizeLatRes = TRUE, curve = c(0,0,1.5,0), edge.color = model_table_simplified_refined$color, nodeLabels = c("","","","","","","","","","","","","","","","","","","","","","","","","","","","","","","","","","","","","","","","","","","","","","","","","","","","","","","","", "RURALITY","","","","LIBERAL","","","","","",  "TRUST","HEALTH\nRISK\nPERCEPTION","ECONOMIC\nRISK\nPERCEPTION","","","","","","","","","BEHAVIORAL\nINTENTION"))

#edge.color = c("darkgray", "blue", "blue", "red", "gray", "blue", "red", "red", "red", "red", "red", "red"), 

legend("bottomright",
       legend = c("negative coefficient", "positive coefficent", "marginally non-significant\ncoefficient (0.05 < p < 0.1)"),
       lty = c(5, 5, 5), col = c("red", "blue", "gray"), lwd = 5, cex = 1.1)


legend("bottomleft",
       legend = c("observed variable", "latent variable"),
        col = c("black"), pch = c(0,1), cex = 1.1)

#summary(separatedtypes, fit.measures = TRUE)


```


```{r DiseasePrevalence_2, echo=FALSE, message=FALSE, warning=FALSE, results=FALSE, include=FALSE}
prev_trustandrisk_total <- '
      # latent variable definitions
      trust =~ Q26_1+Q26_2+Q26_3 + Q26_5+Q26_6+Q26_7+Q26_8
      health_risk =~ Q2_1+Q2_2+Q2_3+Q3_1+Q3_5+Q3_6
      econ_risk =~ Q3_2+Q3_3+Q3_4+Q3_7
      #behavioral breakdown
      indoor_close =~ Q4_1 + Q5_1 + Q6_1 + Q7_1 + Q8_1
      indoor_group =~ Q4_2 + Q5_2 + Q6_2 + Q7_2 + Q8_2
      restaurant =~ Q4_3 + Q5_3 + Q6_3 + Q7_3 + Q8_3
      church =~ Q4_4 + Q5_4 + Q6_4 + Q7_4 + Q8_4
      shop =~ Q4_5 + Q5_5 + Q6_5 + Q7_5 + Q8_5
      appointments =~ Q4_6 + Q5_6 + Q6_6 + Q7_6 + Q8_6
      community =~ Q4_7 + Q5_7 + Q6_7 + Q7_7 + Q8_7
      mask =~  Q5_8 + Q6_8 + Q7_8 + Q8_8
      beh_int =~ indoor_close + indoor_group + restaurant + church + shop + appointments + community + mask 
      
      # regressions
     trust ~ h*ruralbool + femalebool + whitebool + age65plusbool + polVecLiberal + polVecConservative + polVecLibertarian + polVecNonpolitical + polVecOther
     health_risk ~ ruralbool + femalebool + whitebool + age65plusbool + polVecLiberal + polVecConservative + polVecLibertarian + polVecNonpolitical + polVecOther
     econ_risk ~ j*ruralbool + femalebool + whitebool + age65plusbool + polVecLiberal + polVecConservative + polVecLibertarian + polVecNonpolitical + polVecOther
     beh_int ~  econ_risk + health_risk + trust + ruralbool + femalebool + whitebool + age65plusbool + polVecLiberal + polVecConservative + polVecLibertarian + polVecNonpolitical + polVecOther + casetodateper100 
     
    #  T_total := a*f*g + c*e*g #trust in governemnt effect on disease burden
    #  Rh_total := f*g #health risk effect on disease burden
    #  Re_total := e*g #economic risk effect on disease burden
    #  UR_trust := h*T_total #impact of rural on burden via trust in science
    #  UR_econ := j*Re_total #impact of rural on burden via economic risk
    #  UR_total := UR_trust + UR_econ + k #net impact of rural on burden
'
 
prevtotaltandrmodel <- sem(model=prev_trustandrisk_total,data=surveydata, std.ov=T)
```


## Table S3.8: Model 3B - Trust and Risk Perception on Behavioral Intention Summary Table Output

```{r, echo= FALSE, message = FALSE, warning = FALSE, error = FALSE, results = FALSE, include=FALSE}


fit <- summary(prevtotaltandrmodel, fit.measures = TRUE, standardized = TRUE)


model_table <- fit$PE[c(57:104),]
model_table <- as.data.frame(model_table)
model_table$est <- round(as.numeric(model_table$std.all), 3)
model_table$se <- round(as.numeric(model_table$se), 3)
model_table$z <- round(as.numeric(model_table$z), 3)
model_table$pvalue <- round(as.numeric(model_table$pvalue), 3)
model_table <- model_table[,c(1,3,11,7,8,9)]
colnames(model_table) <- c("Latent Variable", "Regression Variable", "std.est", "std.error", "z", "p-value")

model_table_original <- model_table

model_table$`Latent Variable`[2:8] <- " "
model_table$`Latent Variable`[10:17] <- " "
model_table$`Latent Variable`[19:26] <- " "
model_table$`Latent Variable`[28:35] <- " "
model_table$`Latent Variable`[37:48] <- " "

```


```{r, echo= FALSE, message = FALSE, warning = FALSE}

rownames(model_table) <- NULL
library(kableExtra)


kable(model_table, linesep = "")%>%
kable_styling(full_width = F,
                position = "left", font_size = 8) %>%footnote(general = paste("chisq = ", round(fit$FIT[3], 3),  "  aic = ", round(fit$FIT[10], 3), "  rmsea = ", round(fit$FIT[14], 3), sep=""))

# model_table_word <- read_docx(path = "./table_templates/COVID_survey_table_template.docx") %>% body_add_table(head(model_table, n = 33, style = "table_template")) %>% body_add_break()
#   
# caption <- block_caption(paste("chisq = ", round(fit$FIT[3], 3), "  cfi = ",  round(fit$FIT[9], 3), "  aic = ", round(fit$FIT[13], 3), "  rmsea = ", round(fit$FIT[17], 3), sep=""), style = "Normal")
# 
#   body_add_caption(model_table_word,  caption)
  
```
  


```{r, echo= FALSE, message = FALSE, warning = FALSE, include = FALSE}

# print(model_table_word, target = "./tables/Model3_table.docx")
```

## Figure S3.12: Model 3B - Trust and Risk Perception on Behavioral Intention Full Diagram


```{r, echo= FALSE, message = FALSE, warning = FALSE }

obj <- do.call(semPlotModel, list(prevtotaltandrmodel)) # this is more general / not just for Mplus
idx <- apply(obj@Pars[c("lhs", "rhs")], 1, function(i) any(grepl("Q",  i)))
obj@Pars <- obj@Pars[!idx, ]


model_table_diagram <- model_table
model_table_diagram[is.na(model_table_diagram)] = 0
model_table_diagram$color <- ifelse(model_table_diagram$std.est > 0,                  # First test condition
 
       yes = ifelse(model_table_diagram$`p-value` > .05 & model_table_diagram$`p-value` < .1,    # Second test condition
                    yes =  "gray",
                    no = "blue"),
 
       no = ifelse(model_table_diagram$`p-value` > .05 & model_table_diagram$`p-value` < .1,    # Second test condition
                    yes = "gray",
                    no = "red"))

model_table_diagram$sig_binary <- ifelse(model_table_diagram$`p-value` > .1, 1,0)
#model_table_diagram <- model_table_diagram[-c(51:53),]
obj@Pars <- obj@Pars[1:48,]


rownames(model_table_diagram) <- as.numeric(rownames(model_table_diagram))+56

len <- rownames(model_table_diagram[model_table_diagram$sig_binary == 0,])
#obj@Pars <- obj@Pars[rownames(obj@Pars) == rownames(model_table_diagram[model_table_diagram$sig_binary == 0,]), ]   

obj@Pars <- obj@Pars[rownames(obj@Pars) %in% len, ] 


model_table_diagram_refined <- model_table_diagram[model_table_diagram$sig_binary == 0,]


#--create custom layout
ly <- t(matrix(c(9,3,4,3,7,3,11,3,8,3,6,3,5,3,10,3,8,5,3,3,2,3,1,3,10,7,9,7,8,7,7,7,6,7,5,7,4,7,3,7,6,5),2))


semPlot::semPaths(obj, bifactor = "g", fade = FALSE, style = "lisrel", label.cex = .8, nCharNodes = 0,  what = "std", layout=ly, curvePivot = TRUE, edge.label.cex=1, intercepts = FALSE, sizeLat = 8, sizeLat2 =6, rotation = 3, residuals = FALSE, exoVar = FALSE, exoCov = FALSE, label.scale=FALSE, edgeLabels = FALSE, sizeMan = 7, sizeMan2 = 4, allVars = FALSE, mar = c(7,.95,7,.95), thresholds = FALSE, optimizeLatRes = TRUE,  curve = c(0,0,0,0,0,0,0,0,-1,.5,-2,-1,-1,1.3,-1,-1,1,1,1,.4,.5,.8,.8,0,0,0,0,0,0,0), edge.color = model_table_diagram_refined$color, nodeLabels = c("", "", "", "", "", "", "", "", "", "", "", "", "", "", "", "", "", "", "", "", "", "", "", "", "", "", "", "", "", "", "", "", "", "", "", "", "", "", "", "", "", "", "", "", "", "", "", "", "", "", "", "", "", "", "", "",    "WHITE", "NONPOL", "LIBERTRN", "RURAL","AGE 65+", "LIB", "CONS", "FEMALE", "",  "CASES", "TRUST", "HEALTH\nRISK", "ECONOMIC\nRISK", "INDOOR\nCLOSE", "INDOOR\nGROUP", "RESTRNTS", "CHURCH", "SHOPPING", "APPTS", "COMMUNITY", "MASK",  "BEHAVIOR", "", "", "" ))


legend("bottomright",
       legend = c("negative coefficient", "positive coefficent", "marginally non-significant\ncoefficient (0.05 < p < 0.1)"),
       lty = c(5, 5, 5), col = c("red", "blue", "gray"), lwd = 5, cex = 1.1)


legend("bottomleft",
       legend = c("observed variable", "latent variable"),
        col = c("black"), pch = c(0,1), cex = 1.1)

#summary(separatedtypes, fit.measures = TRUE)
```


## Figure S3.13: Model 3B - Trust and Risk Perception on Behavioral Intention Simplified Diagram

```{r, echo= FALSE, echo=FALSE, message=FALSE, warning=FALSE, results=FALSE }


obj <- do.call(semPlotModel, list(prevtotaltandrmodel)) # this is more general / not just for Mplus


model_table_simplified <- model_table_original[c("65", "74","83", "92", "93", "94", "95",  "104"),]
model_table_simplified$color <- ifelse(model_table_simplified$std.est > 0,                  # First test condition
 
       yes = ifelse(model_table_simplified$`p-value` > .05,    # Second test condition
                    yes =  "gray",
                    no = "blue"),
 
       no = ifelse(model_table_simplified$`p-value` > .05,    # Second test condition
                    yes = "gray",
                    no = "red"))

model_table_simplified$sig_binary <- ifelse(model_table_simplified$`p-value` > .1, 1,0)
model_table_simplified_refined <- model_table_simplified[model_table_simplified$sig_binary == 0,]


obj@Pars <- obj@Pars[rownames(model_table_simplified_refined),]
ly <- t(matrix(c(4,3,4,2,1,2,2,3,2,1,3,2), 2))


semPlot::semPaths(obj, bifactor = "g", fade = FALSE, style = "lisrel", label.cex = .8, nCharNodes = 0,  what = "std", layout=ly, curvePivot = TRUE, edge.label.cex=1, intercepts = FALSE, sizeLat = 14, sizeLat2 =8, rotation = 3, residuals = FALSE, exoVar = FALSE, exoCov = FALSE, label.scale=FALSE, layoutSplit = TRUE, edgeLabels = TRUE, sizeMan = 10, sizeMan2 = 5, allVars = FALSE, mar = c(7,3,7,3), thresholds = FALSE, optimizeLatRes = TRUE, curve = c(-.7,1,0,0,0,0), edge.color = model_table_simplified_refined$color, nodeLabels = c("","","","","","","","","","","","","","","","","","","","","","","","","","","","","","","","","","","","","","","","","","","","","","","","","","","","","","","","","RURALITY","","","","LIBERAL","","","","","CASES","TRUST","HEALTH\nRISK\nPERCEPTION","ECONOMIC\nRISK\nPERCEPTION","","","","","","","","","BEHAVIORAL\nINTENTION"))

#edge.color = c("darkgray", "blue", "blue", "red", "gray", "blue", "red", "red", "red", "red", "red", "red"), 

legend("bottomright",
       legend = c("negative coefficient", "positive coefficent", "marginally non-significant\ncoefficient (0.05 < p < 0.1)"),
       lty = c(5, 5, 5), col = c("red", "blue", "gray"), lwd = 5, cex = 1.1)


legend("bottomleft",
       legend = c("observed variable", "latent variable"),
        col = c("black"), pch = c(0,1), cex = 1.1)

#summary(separatedtypes, fit.measures = TRUE)


```

\newpage

## Table S3.9: Comparing Model Set A

```{r modelA_comparison, echo=FALSE} 
model1A <- caseburden
model2A <- caseburdenrtot
model3A <- caseburdentandr

lavTestLRT(model1A,model2A)
lavTestLRT(model1A,model3A)
```
\newpage

## Table S3.10: Comparing Model Set B

```{r modelB_comparison, echo=FALSE} 
model1B <- prevtotalmodel
model2B <- prevtotalrtmodel
model3B <- prevtotaltandrmodel

lavTestLRT(model1B,model2B)
lavTestLRT(model1B,model3B)
```
````
