## Supplementary material for "Effects of trust, risk perception, and health behavior on COVID-19 disease burden: Evidence from a multi-state US survey": S3. Compiled R markdown

### Supplemental Appendix

#### Supplemental Appendix Overview

This supplemental appendix provides full analysis outputs, associated reproducible code, and supportive data. As noted in the associated paper, our analysis describes relationships between social cognitive measures (collected from our COVID-19 survey, conducted in the Fall of 2020) and COVID-19 cases (aggregated by the New York Times - http://github.com/nytimes/covid19).

##### Data Collection and Transformation

Our structural equation models rely upon the following datasets:

1. COVID-19 daily case data, at a county level. We use aggregate case data that is assembled by the New York Times, and is exposed via their github.com repository (http://github.com/nytimes/covid19). This dataset is within our /data folder, named "COVID_data.csv".

2. COVID-19 survey data. We conducted a COVID-19 health perceptions survey in the Fall of 2020 (n = 1035). This de-identified survey dataset is within our /data folder, named "COVID_survey.csv".

From a transformation perspective, several additional fields are generated:

1. Data is reduced to only cases for Idaho, Texas and Vermont;
2. COVID-19 cases per county are grouped based on 1) cumulative cases between June 1, 2020 and April 30th, 2021, as well as 2) cumulative cases 15 days before and after survey completion.
3. In order to assign rurality, survey responses were grouped into urban and rural classifications, based on the United States Department of Agriculture's Rural/Urban Commuting Areas (RUCA)

#### Conceptual Models Overview

As note in our associated paper, we evaluated 6 (six) models. Please note the following variable naming conventions: 1) 'Health Risk' and 'Economic Risk' refer to health risk perception and economic risk perception, respectfully. 2) "Behavior" refers to both current behavior (at the time of survey completion) as well as behavioral intention. Shorter terms are used for brevity in diagram notation.

**Model Set A:** Behavioral intentions as an influence on disease burden. Here we consider the disease burden as cumulative cases across a large window of time (June 2020 to April 2021) to capture a complete view of disease impact.

- Model 1A: Trust effects on economic risk perception and health risk perception, to behavioral intention, and then to disease burden.
- Model 2A: Economic risk perception and health risk perception effects on trust, which impact behavioral intention, and then disease burden.
- Model 3A: Trust, economic risk perception and health risk perception - all impacting behavioral intention directly, and then to disease burden.

**Model Set B:** Disease burden as an influence on behavioral intention. Here we consider a snapshot of the cases in the two weeks prior to the survey response as a measure of the local disease burden and consider its impact on the expressed behavioral intention.

- Model 1B: Trust effects on economic risk perception and health risk perception, and then behavioral intention.
- Model 2B: Economic risk perception and health risk perception effects on trust, which impact behavioral intention.
- Model 3B: Trust, economic risk perception and health risk perception - all impacting behavioral intention directly.

#### Table S3.1: Survey Demographics

| | ID Pct | TX Pct | VT Pct | Total Pct |
|---|---|---|---|---|
| **Age** | | | | |
| Under 25 | 3.0 | 4.4 | 3.4 | 3.3 |
| 25-35 | 10.9 | 12.6 | 11.7 | 11.4 |
| 35-45 | 15.1 | 17.0 | 11.1 | 13.9 |
| 45-55 | 15.1 | 16.3 | 13.4 | 14.6 |
| 55-65 | 20.6 | 19.3 | 25.1 | 22.1 |
| 65-75 | 25.7 | 17.8 | 26.8 | 25.0 |
| 75+ | 9.6 | 12.6 | 8.5 | 9.6 |
| **Educational Attainment** | | | | |
| Did not graduate HS | 0.4 | 0.7 | 0.3 | 0.4 |
| HS diploma or equivalent | 7.5 | 13.9 | 7.5 | 8.4 |
| Some college | 19.6 | 12.4 | 11.7 | 15.7 |
| Associate's degree | 11.7 | 9.5 | 5.3 | 9.1 |
| College graduate or higher | 60.9 | 63.5 | 75.1 | 66.4 |
| **Ethnicity** | | | | |
| Hispanic | 4.3 | 11.1 | 0.6 | 3.9 |
| Non-Hispanic | 95.7 | 88.9 | 99.4 | 96.1 |
| **Gender** | | | | |
| Male | 42.0 | 48.5 | 45.7 | 44.2 |
| Female | 57.4 | 50.7 | 53.8 | 55.2 |
| Not Identified | 0.6 | 0.7 | 0.6 | 0.6 |
| **Income Bracket** | | | | |
| <$25,000 | 7.0 | 8.7 | 6.7 | 7.1 |
| $25,000-$49,999 | 21.1 | 15.7 | 17.0 | 18.9 |
| $50,000-$74,999 | 24.8 | 19.7 | 20.2 | 22.4 |
| >$74,999 | 47.2 | 55.9 | 56.0 | 51.6 |
| **Politics** | | | | |
| Moderate | 28.8 | 35.6 | 28.5 | 29.6 |
| Liberal | 19.0 | 15.6 | 46.6 | 28.5 |
| Conservative | 37.3 | 34.1 | 13.3 | 28.2 |
| Libertarian | 4.1 | 4.4 | 2.5 | 3.6 |
| Nonpolitical | 8.6 | 7.4 | 7.1 | 7.9 |
| Other | 2.2 | 3.0 | 2.0 | 2.2 |
| **Race** | | | | |
| No Answer | 8.0 | 9.0 | 3.8 | 6.7 |
| American Indian or Alaska Native | 0.2 | 0.7 | 0.0 | 0.2 |
| Multi-race | 3.8 | 3.5 | 0.8 | 2.7 |
| Asian or Asian-American | 1.1 | 4.2 | 1.1 | 1.5 |
| Black or African-American | 0.2 | 6.9 | 0.8 | 1.4 |
| Native Hawaiian or Other Pacific Islander | 0.4 | 0.0 | 0.0 | 0.2 |
| White | 83.5 | 72.9 | 91.8 | 84.9 |

*(continued)*

| | ID Pct | TX Pct | VT Pct | Total Pct |
|---|---|---|---|---|
| Other | 2.9 | 2.8 | 1.6 | 2.4 |
| **Relationship Status** | | | | |
| Single, never married | 8.4 | 16.3 | 16.5 | 12.4 |
| Married or domestic partnership | 72.0 | 67.4 | 61.7 | 67.7 |
| Widowed | 6.3 | 7.4 | 5.3 | 6.1 |
| Divorced | 12.4 | 6.7 | 15.1 | 12.6 |
| Separated | 0.8 | 2.2 | 1.4 | 1.2 |
| **Religion** | | | | |
| Evangelical | 19.5 | 29.1 | 7.7 | 16.6 |
| Catholic | 12.5 | 21.6 | 19.8 | 16.4 |
| Mainline Protestant | 12.7 | 20.9 | 16.3 | 15.2 |
| Latter-Day Saints | 21.4 | 0.0 | 1.8 | 11.3 |
| Jewish | 0.4 | 0.0 | 3.8 | 1.6 |
| Islam | 0.0 | 2.2 | 0.3 | 0.4 |
| Buddhist | 1.5 | 0.0 | 2.4 | 1.6 |
| Atheist | 6.4 | 3.7 | 18.3 | 10.3 |
| Agnostic | 12.7 | 9.0 | 16.9 | 13.7 |
| Other | 12.7 | 13.4 | 12.7 | 12.8 |
| **Rurality** | | | | |
| Rural | 36.3 | 27.8 | 55.6 | 41.9 |
| Urban | 63.7 | 72.2 | 44.4 | 58.1 |

*Note:*

Totals: ID: n = 526 TX: n = 143 VT: n = 365

Age: chi sq. = 12.48, df = 12, p-value = 0.408

Educational Attainment: chi sq. = 31.37, df = 8, p-value = 0

Ethnicity: chi sq. = 29.45, df = 2, p-value = 0

Gender: chi sq. = 2.4, df = 4, p-value = 0.663

Income Bracket: chi sq. = 8.77, df = 6, p-value = 0.187

Politics: chi sq. = 113.04, df = 10, p-value = 0

Race: chi sq. = 71.64, df = 14, p-value = 0

Relationship Status: chi sq. = 24.3, df = 8, p-value = 0.002

Religion: chi sq. = 201.83, df = 18, p-value = 0

Rurality: chi sq. = 46.74, df = 2, p-value = 0

#### Table S3.2: RUCA Code Descriptions

| Primary RUCA Codes | Description |
|---:|---|
| 1 | Metropolitan area core: primary flow within an urbanized area (UA) |
| 2 | Metropolitan area high commuting: primary flow 30% or more to a UA |
| 3 | Metropolitan area low commuting: primary flow 10% to 30% to a UA |
| 4 | Micropolitan area core: primary flow within an Urban Cluster of 10,000 to 49,999 (large UC) |
| 5 | Micropolitan high commuting: primary flow 30% or more to a large UC |
| 6 | Micropolitan low commuting: primary flow 10% to 30% to a large UC |
| 7 | Small town core: primary flow within an Urban Cluster of 2,500 to 9,999 (small UC) |
| 8 | Small town high commuting: primary flow 30% or more to a small UC |
| 9 | Small town low commuting: primary flow 10% to 30% to a small UC |
| 10 | Rural areas: primary flow to a tract outside a UA or UC |
| 99 | Not coded: Census tract has zero population and no rural-urban identifier information |

#### Figure S3.1: Conceptual Models

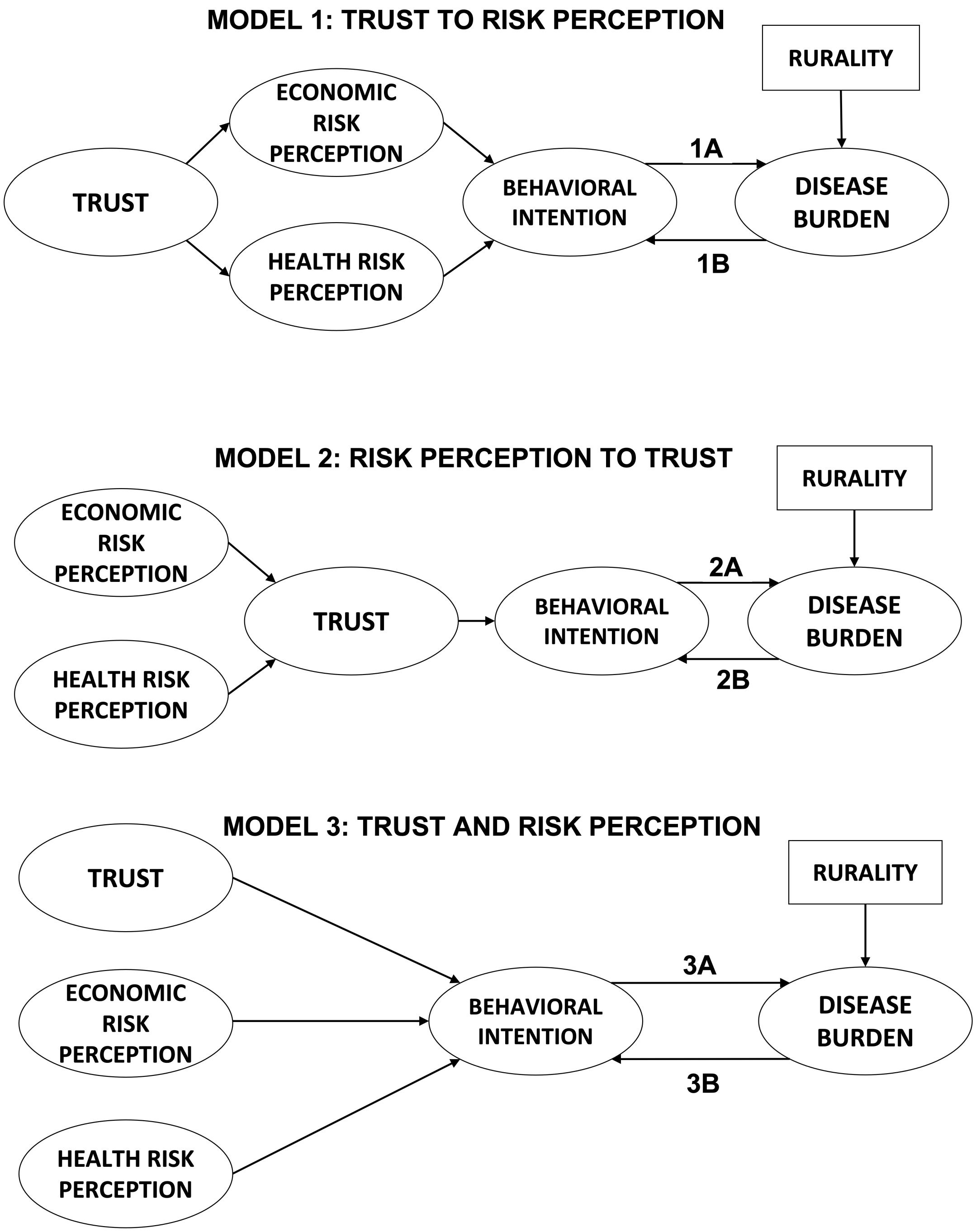

#### Figure S3.2: Behavioral Intention Latent Variable Input

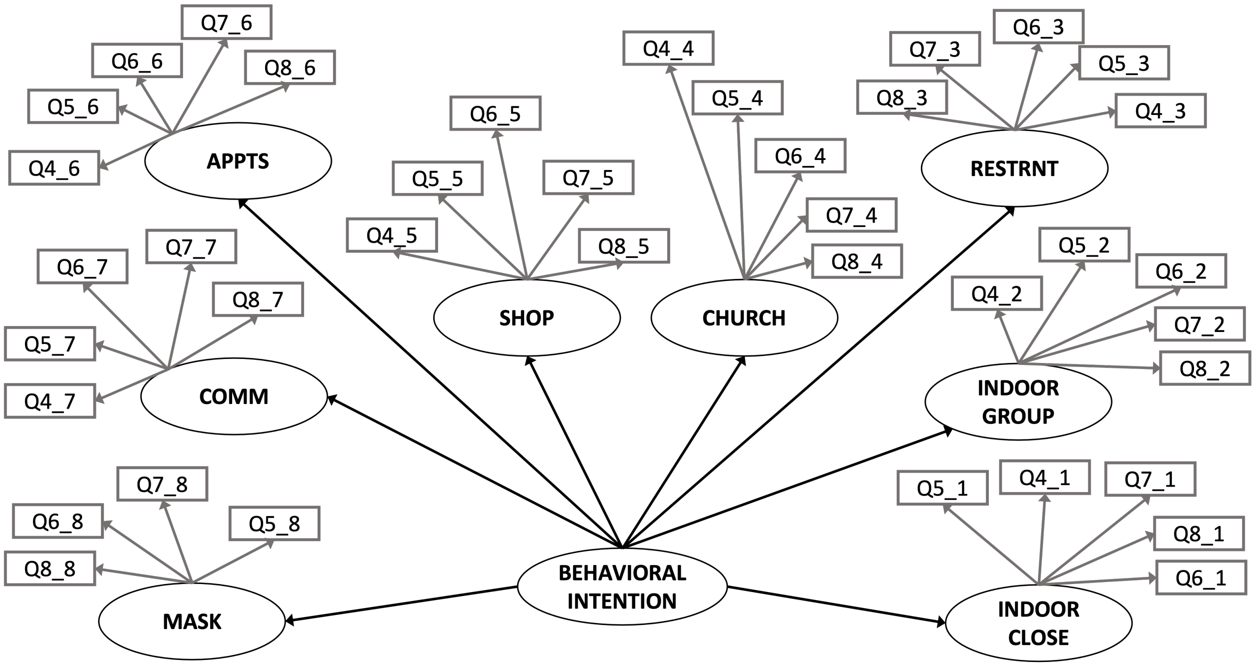

#### Figure S3.3: Trust, Health Risk Perception, and Economic Risk Perception Latent Variable Input

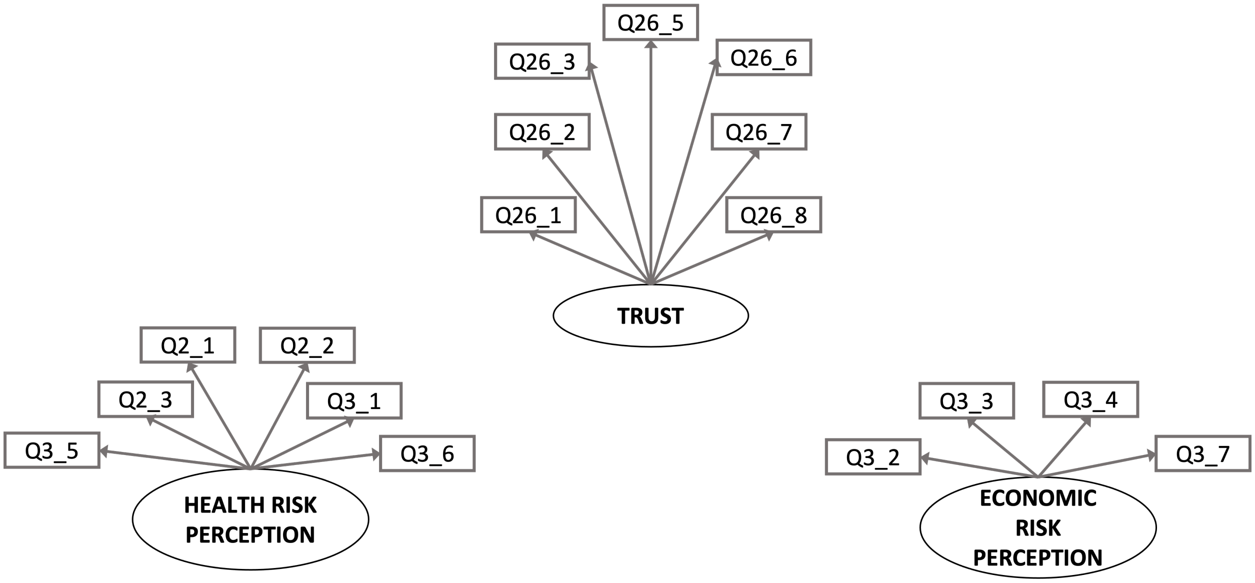

#### Table S3.3: Model 1A - Trust to Risk Perception on Disease Burden Summary Output Table

| Latent Variable | Regression Variable | std.est | std.error | z | p-value |
|---|---|---|---|---|---|
| beh_int | indoor_close | 0.6311948 | 0.000 | NA | NA |
| | indoor_group | 0.8581162 | 1.244 | 2.393 | 0.017 |
| | restaurant | 0.8622779 | 0.893 | 2.144 | 0.032 |
| | church | 0.8192714 | 2.651 | 2.576 | 0.010 |
| | shop | 0.4820082 | 0.396 | 1.695 | 0.090 |
| | appointments | 0.6341776 | 0.531 | 1.769 | 0.077 |
| | community | 0.8581193 | 0.906 | 2.175 | 0.030 |
| | mask | 0.6126211 | 3.225 | 2.590 | 0.010 |
| trust | ruralbool | -0.0523744 | 0.027 | -1.707 | 0.088 |
| | femalebool | 0.0471461 | 0.027 | 1.529 | 0.126 |
| | whitebool | 0.1198891 | 0.047 | 3.747 | 0.000 |
| | age65plusbool | 0.0355694 | 0.029 | 1.153 | 0.249 |
| | polVecLiberal | 0.1269436 | 0.036 | 3.469 | 0.001 |
| | polVecConservative | -0.4791799 | 0.049 | -9.551 | 0.000 |
| | polVecLibertarian | -0.2718832 | 0.087 | -7.265 | 0.000 |
| | polVecNonpolitical | -0.1254426 | 0.058 | -3.671 | 0.000 |
| | polVecOther | -0.0278693 | 0.099 | -0.899 | 0.369 |
| health_risk | trust | 0.6029439 | 0.112 | 9.992 | 0.000 |
| | ruralbool | 0.0176077 | 0.046 | 0.630 | 0.529 |
| | femalebool | 0.1073648 | 0.046 | 3.804 | 0.000 |
| | whitebool | -0.1224351 | 0.078 | -4.271 | 0.000 |
| | age65plusbool | 0.2027356 | 0.050 | 7.098 | 0.000 |
| | polVecLiberal | 0.0814432 | 0.060 | 2.482 | 0.013 |
| | polVecConservative | -0.0484865 | 0.068 | -1.293 | 0.196 |
| | polVecLibertarian | -0.0595328 | 0.133 | -1.933 | 0.053 |
| | polVecNonpolitical | 0.0277186 | 0.096 | 0.909 | 0.363 |
| | polVecOther | 0.0430336 | 0.169 | 1.518 | 0.129 |
| econ_risk | trust | 0.2311012 | 0.052 | 3.845 | 0.000 |
| | ruralbool | -0.0654408 | 0.030 | -1.667 | 0.096 |
| | femalebool | 0.0838719 | 0.031 | 2.105 | 0.035 |
| | whitebool | -0.0647868 | 0.051 | -1.617 | 0.106 |
| | age65plusbool | 0.0037188 | 0.032 | 0.095 | 0.924 |
| | polVecLiberal | 0.0506782 | 0.039 | 1.109 | 0.267 |
| | polVecConservative | -0.1508310 | 0.046 | -2.796 | 0.005 |
| | polVecLibertarian | 0.0995256 | 0.088 | 2.276 | 0.023 |
| | polVecNonpolitical | -0.0077235 | 0.062 | -0.183 | 0.855 |
| | polVecOther | 0.0212453 | 0.109 | 0.540 | 0.589 |
| beh_int | econ_risk | 0.0577769 | 0.006 | 1.573 | 0.116 |
| | health_risk | -0.7020007 | 0.020 | -2.596 | 0.009 |
| | ruralbool | 0.0063131 | 0.003 | 0.268 | 0.788 |
| | femalebool | 0.0747554 | 0.005 | 2.007 | 0.045 |
| | whitebool | -0.0367731 | 0.006 | -1.339 | 0.180 |
| | age65plusbool | -0.0095526 | 0.003 | -0.386 | 0.699 |
| | polVecLiberal | -0.0947068 | 0.006 | -2.083 | 0.037 |
| | polVecConservative | 0.2347362 | 0.013 | 2.484 | 0.013 |
| | polVecLibertarian | 0.0597318 | 0.011 | 1.759 | 0.079 |
| | polVecNonpolitical | 0.0256736 | 0.006 | 0.949 | 0.343 |
| | polVecOther | 0.0098239 | 0.011 | 0.410 | 0.682 |
| cumcasesper100 | beh_int | 0.2637917 | 1.743 | 2.482 | 0.013 |
| | ruralbool | -0.2728009 | 0.065 | -8.442 | 0.000 |
| Trust | | -0.1116543 | 0.042 | -6.093 | 0.000 |
| Health_risk | | -0.1851820 | 0.031 | -7.374 | 0.000 |
| Rural_via_trust | | 0.0058478 | 0.007 | 1.668 | 0.095 |

*Note:*

chisq = 10470.078 aic = 98774.041 rmsea = 0.071

#### Figure S3.4: Model 1A - Trust to Risk Perception on Disease Burden Full Diagram

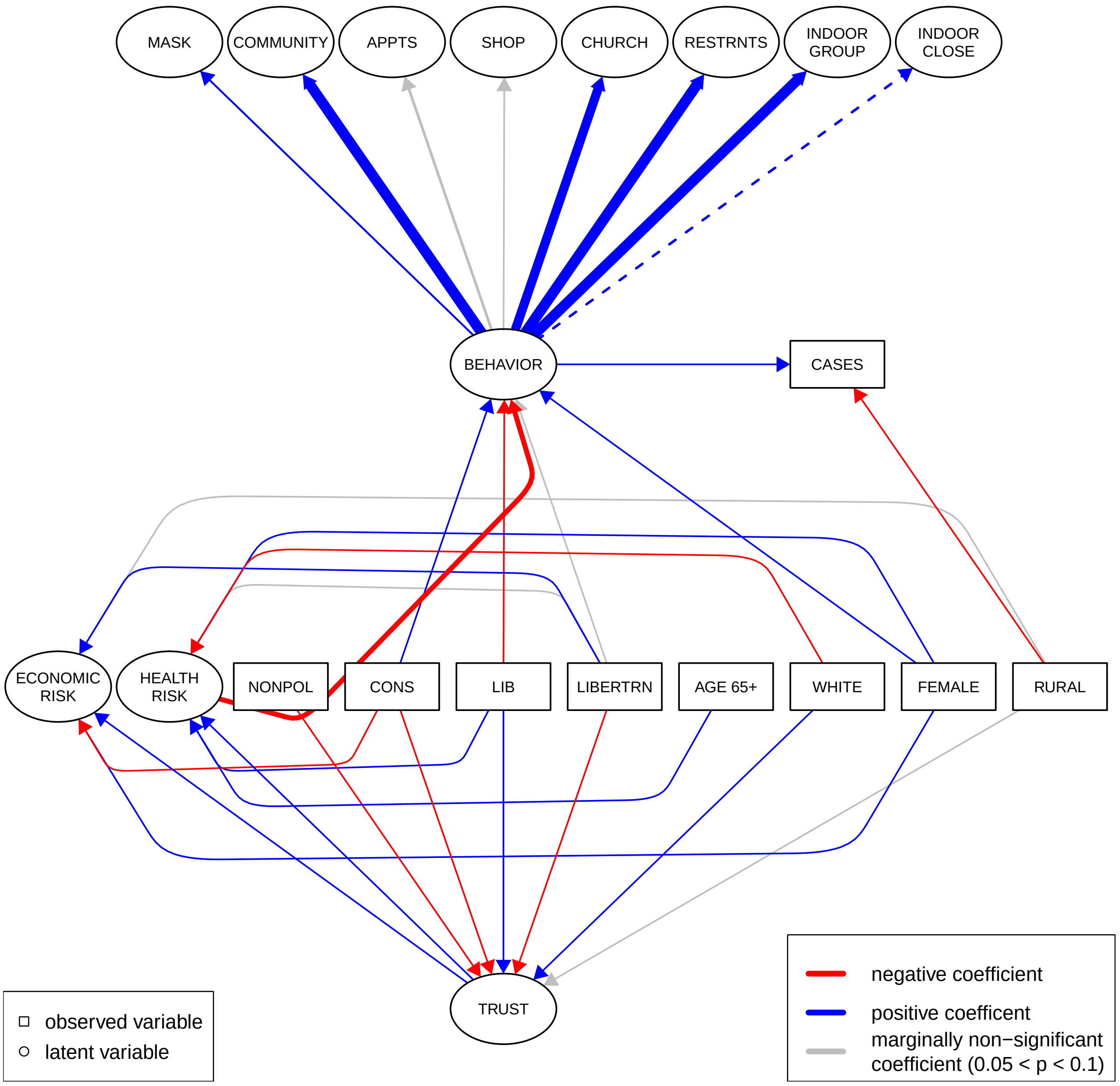

#### Figure S3.5: Model 1A - Trust to Risk Perception on Disease Burden Simplified Diagram

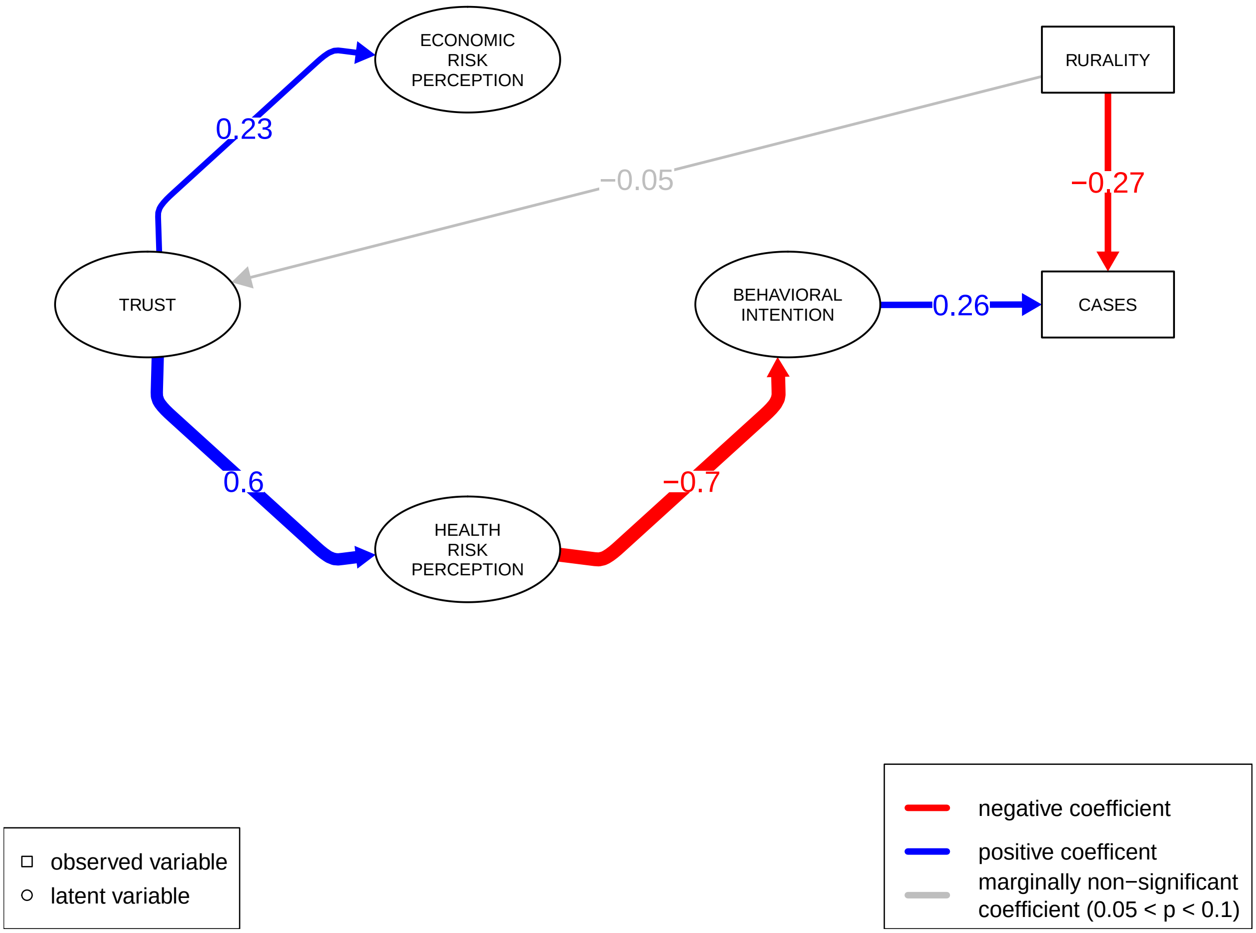

#### Table S3.4: Model 2A Risk Perception to Trust on Disease Burden Summary Table Output

| Latent Variable | Regression Variable | std.est | std.error | z | p-value |
|---|---|---|---|---|---|
| beh__int | indoor__close | 0.6154574 | 0.000 | NA | NA |
| | indoor__group | 0.8636016 | 1.274 | 2.401 | 0.016 |
| | restaurant | 0.8564154 | 0.905 | 2.150 | 0.032 |
| | church | 0.8192486 | 2.689 | 2.586 | 0.010 |
| | shop | 0.4676550 | 0.392 | 1.701 | 0.089 |
| | appointments | 0.6266757 | 0.538 | 1.783 | 0.075 |
| | community | 0.8586784 | 0.924 | 2.181 | 0.029 |
| | mask | 0.6163858 | 3.295 | 2.599 | 0.009 |
| trust | ruralbool | -0.0461582 | 0.023 | -1.733 | 0.083 |
| | femalebool | -0.0271581 | 0.024 | -1.003 | 0.316 |
| | whitebool | 0.1472786 | 0.041 | 5.111 | 0.000 |
| | age65plusbool | -0.0898629 | 0.026 | -3.167 | 0.002 |
| | polVecLiberal | 0.0398757 | 0.030 | 1.276 | 0.202 |
| | polVecConservative | -0.2976379 | 0.039 | -7.371 | 0.000 |
| | polVecLibertarian | -0.1452722 | 0.069 | -4.790 | 0.000 |
| | polVecNonpolitical | -0.0991984 | 0.049 | -3.371 | 0.001 |
| | polVecOther | -0.0421418 | 0.085 | -1.566 | 0.117 |
| | health__risk | 0.5658121 | 0.029 | 10.215 | 0.000 |
| | econ__risk | -0.0276393 | 0.037 | -0.874 | 0.382 |
| health__risk | ruralbool | -0.0145403 | 0.052 | -0.461 | 0.645 |
| | femalebool | 0.1362374 | 0.052 | 4.264 | 0.000 |
| | whitebool | -0.0498518 | 0.087 | -1.562 | 0.118 |
| | age65plusbool | 0.2228255 | 0.056 | 6.896 | 0.000 |
| | polVecLiberal | 0.1577016 | 0.067 | 4.276 | 0.000 |
| | polVecConservative | -0.3373801 | 0.069 | -8.933 | 0.000 |
| | polVecLibertarian | -0.2231336 | 0.143 | -6.724 | 0.000 |
| | polVecNonpolitical | -0.0483513 | 0.107 | -1.416 | 0.157 |
| | polVecOther | 0.0255556 | 0.191 | 0.797 | 0.426 |
| econ__risk | ruralbool | -0.0787981 | 0.029 | -2.003 | 0.045 |
| | femalebool | 0.0915440 | 0.030 | 2.295 | 0.022 |
| | whitebool | -0.0353964 | 0.048 | -0.906 | 0.365 |
| | age65plusbool | 0.0088206 | 0.031 | 0.227 | 0.821 |
| | polVecLiberal | 0.0775195 | 0.037 | 1.706 | 0.088 |
| | polVecConservative | -0.2533050 | 0.043 | -4.861 | 0.000 |
| | polVecLibertarian | 0.0438878 | 0.078 | 1.091 | 0.275 |
| | polVecNonpolitical | -0.0378155 | 0.059 | -0.905 | 0.365 |
| | polVecOther | 0.0133654 | 0.105 | 0.342 | 0.733 |
| beh__int | trust | -0.6246913 | 0.034 | -2.550 | 0.011 |
| | ruralbool | -0.0202233 | 0.003 | -0.752 | 0.452 |
| | femalebool | 0.0147160 | 0.003 | 0.555 | 0.579 |
| | whitebool | 0.0706311 | 0.007 | 1.878 | 0.060 |
| | age65plusbool | -0.1449152 | 0.008 | -2.376 | 0.017 |
| | polVecLiberal | -0.1211492 | 0.007 | -2.198 | 0.028 |
| | polVecConservative | 0.1573775 | 0.009 | 2.274 | 0.023 |
| | polVecLibertarian | 0.0496342 | 0.011 | 1.456 | 0.145 |
| | polVecNonpolitical | -0.0209865 | 0.007 | -0.718 | 0.473 |
| | polVecOther | -0.0251222 | 0.012 | -0.904 | 0.366 |
| cumcasesper100 | beh__int | 0.2872415 | 1.915 | 2.511 | 0.012 |
| | ruralbool | -0.2734161 | 0.065 | -8.518 | 0.000 |

*Note:*

chisq = 10648.493 aic = 98950.456 rmsea = 0.072

#### Figure S3.6: Model 2A - Risk Perception to Trust on Disease Burden Full Diagram

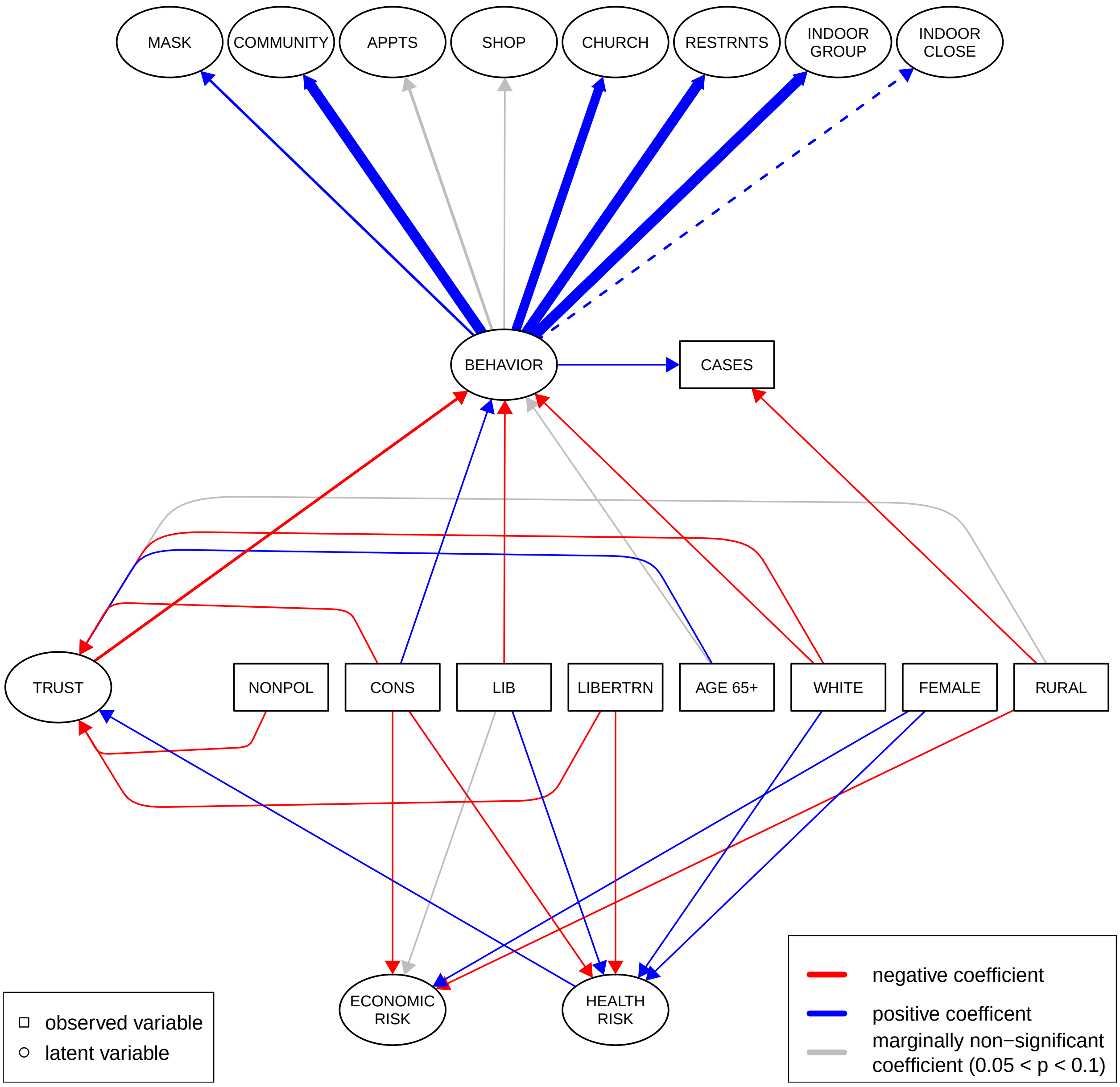

#### Figure S3.7: Model 2A - Risk Perception to Trust on Disease Burden Simplified Diagram

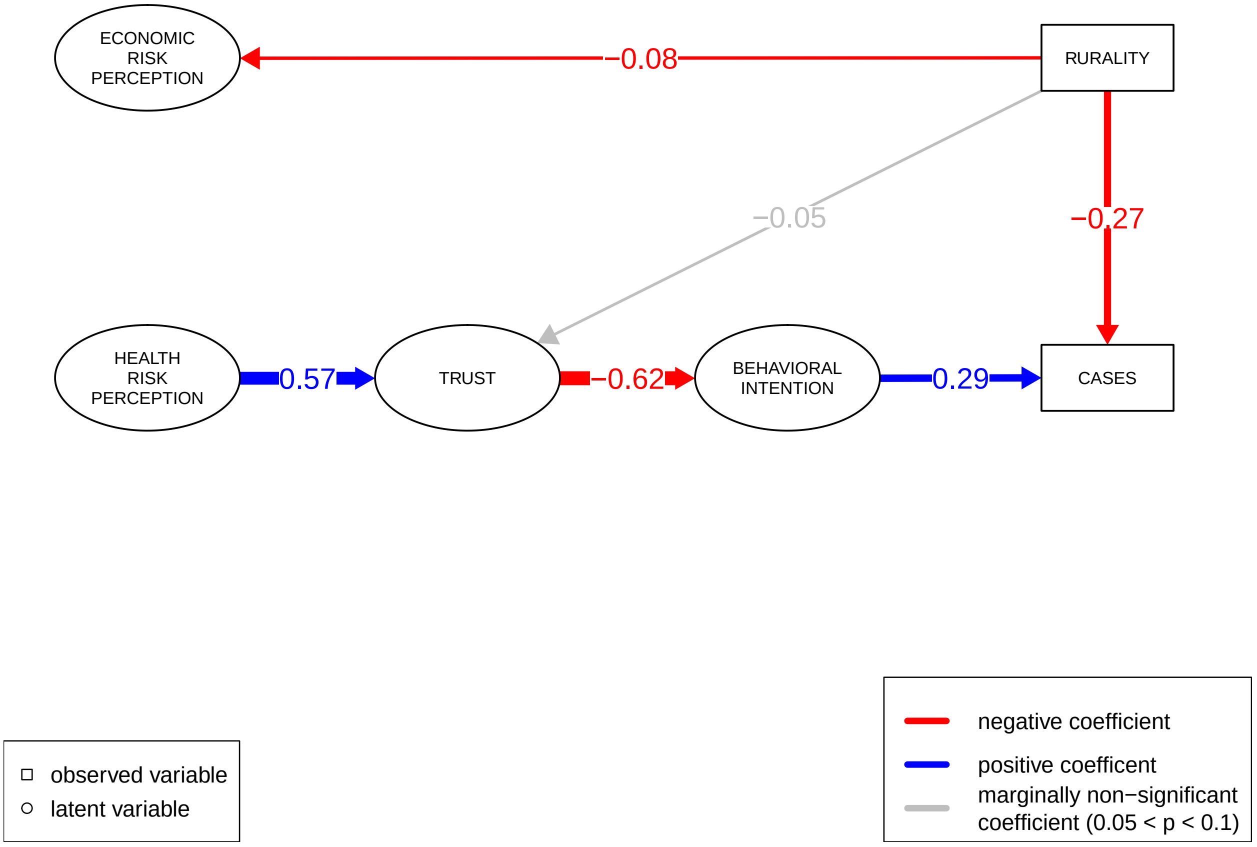

#### Table S3.5: Model 3A Trust and Risk Perception on Disease Burden Summary Table Output

| Latent Variable | Regression Variable | std.est | std.error | z | p-value |
|---|---|---|---|---|---|
| beh__int | indoor__close | 0.6130415 | 0.000 | NA | NA |
| | indoor__group | 0.8528264 | 1.275 | 2.350 | 0.019 |
| | restaurant | 0.8488089 | 0.911 | 2.101 | 0.036 |
| | church | 0.8078437 | 2.697 | 2.535 | 0.011 |
| | shop | 0.4606525 | 0.397 | 1.673 | 0.094 |
| | appointments | 0.6139118 | 0.538 | 1.740 | 0.082 |
| | community | 0.8497749 | 0.928 | 2.130 | 0.033 |
| | mask | 0.6048508 | 3.324 | 2.549 | 0.011 |
| trust | ruralbool | -0.0504009 | 0.030 | -1.633 | 0.103 |
| | femalebool | 0.0450828 | 0.030 | 1.452 | 0.146 |
| | whitebool | 0.1237901 | 0.050 | 3.856 | 0.000 |
| | age65plusbool | 0.0380944 | 0.032 | 1.225 | 0.221 |
| | polVecLiberal | 0.1261649 | 0.039 | 3.441 | 0.001 |
| | polVecConservative | -0.4703305 | 0.050 | -9.834 | 0.000 |
| | polVecLibertarian | -0.2694991 | 0.092 | -7.355 | 0.000 |
| | polVecNonpolitical | -0.1249602 | 0.062 | -3.653 | 0.000 |
| | polVecOther | -0.0295334 | 0.108 | -0.945 | 0.344 |
| health__risk | ruralbool | -0.0129802 | 0.053 | -0.410 | 0.682 |
| | femalebool | 0.1338093 | 0.054 | 4.184 | 0.000 |
| | whitebool | -0.0507499 | 0.088 | -1.587 | 0.113 |
| | age65plusbool | 0.2272469 | 0.058 | 7.029 | 0.000 |
| | polVecLiberal | 0.1562608 | 0.068 | 4.232 | 0.000 |
| | polVecConservative | -0.3317360 | 0.070 | -8.800 | 0.000 |
| | polVecLibertarian | -0.2203727 | 0.146 | -6.641 | 0.000 |
| | polVecNonpolitical | -0.0452897 | 0.109 | -1.324 | 0.185 |
| | polVecOther | 0.0289099 | 0.195 | 0.899 | 0.369 |
| econ__risk | ruralbool | -0.0789176 | 0.029 | -2.007 | 0.045 |
| | femalebool | 0.0914534 | 0.029 | 2.294 | 0.022 |
| | whitebool | -0.0353031 | 0.048 | -0.904 | 0.366 |
| | age65plusbool | 0.0088372 | 0.031 | 0.227 | 0.820 |
| | polVecLiberal | 0.0774280 | 0.037 | 1.705 | 0.088 |
| | polVecConservative | -0.2530092 | 0.043 | -4.853 | 0.000 |
| | polVecLibertarian | 0.0440567 | 0.078 | 1.096 | 0.273 |
| | polVecNonpolitical | -0.0379060 | 0.059 | -0.908 | 0.364 |
| | polVecOther | 0.0132560 | 0.105 | 0.339 | 0.735 |
| beh__int | econ__risk | 0.0570876 | 0.006 | 1.567 | 0.117 |
| | health__risk | -0.5859893 | 0.016 | -2.550 | 0.011 |
| | trust | -0.3008314 | 0.015 | -2.450 | 0.014 |
| | ruralbool | -0.0056390 | 0.003 | -0.237 | 0.813 |
| | femalebool | 0.0709680 | 0.004 | 1.935 | 0.053 |
| | whitebool | 0.0051738 | 0.005 | 0.213 | 0.831 |
| | age65plusbool | -0.0295435 | 0.003 | -1.082 | 0.279 |
| | polVecLiberal | -0.0830591 | 0.006 | -1.941 | 0.052 |
| | polVecConservative | 0.1538314 | 0.009 | 2.243 | 0.025 |
| | polVecLibertarian | 0.0154552 | 0.008 | 0.563 | 0.573 |
| | polVecNonpolitical | -0.0024890 | 0.006 | -0.096 | 0.923 |
| | polVecOther | -0.0005295 | 0.010 | -0.022 | 0.982 |
| cumcasesper100 | beh__int | 0.2600091 | 1.816 | 2.443 | 0.015 |
| cumcasesper100 | ruralbool | -0.2737575 | 0.065 | -8.461 | 0.000 |

*Note:*

chisq = 10629.99 aic = 98931.952 rmsea = 0.072

#### Figure S3.8: Model 3A - Trust and Risk Perception on Disease Burden Full Diagram

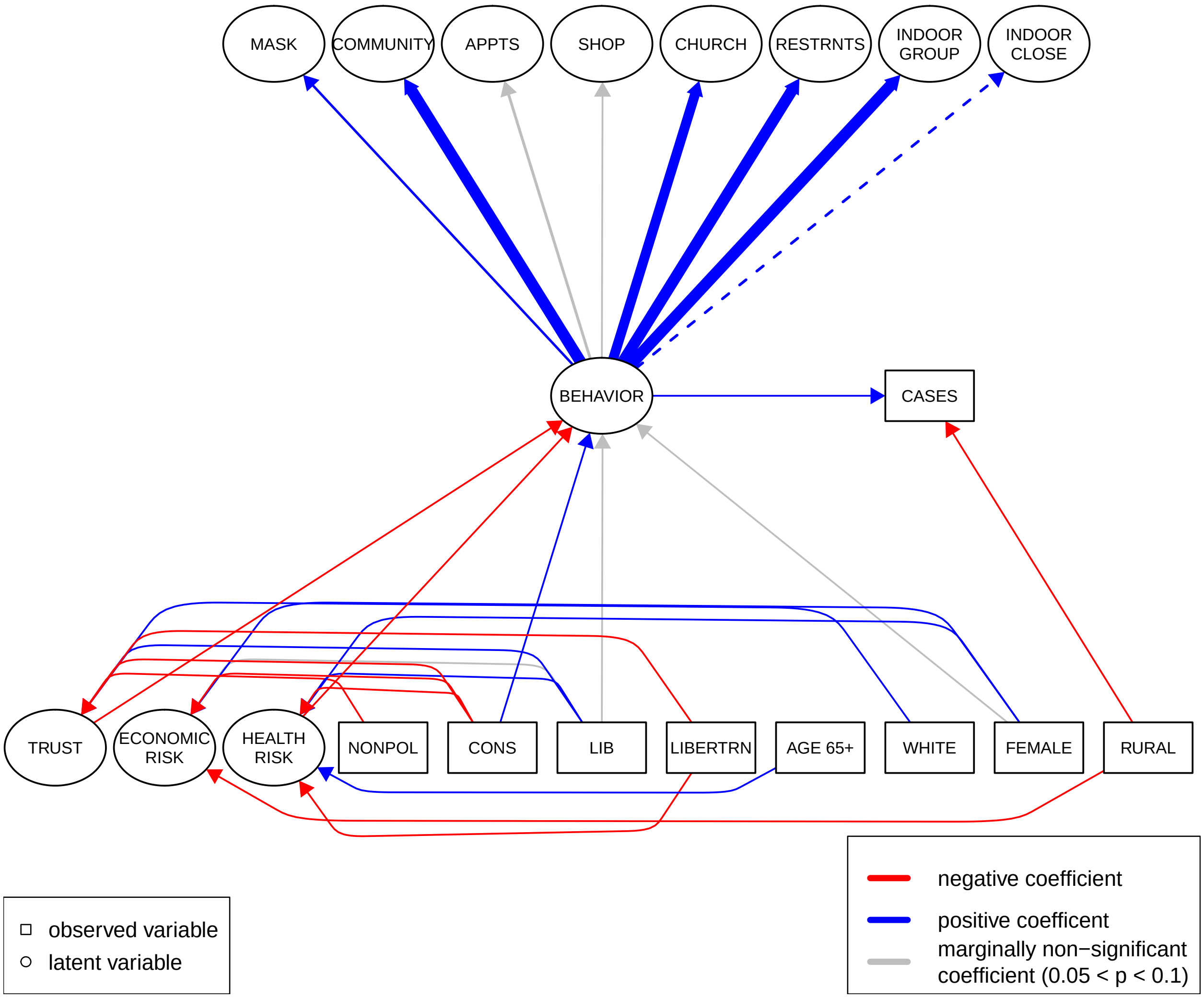

#### Figure S3.9: Model 3A - Trust and Risk Perception on Disease Burden Simplified Diagram

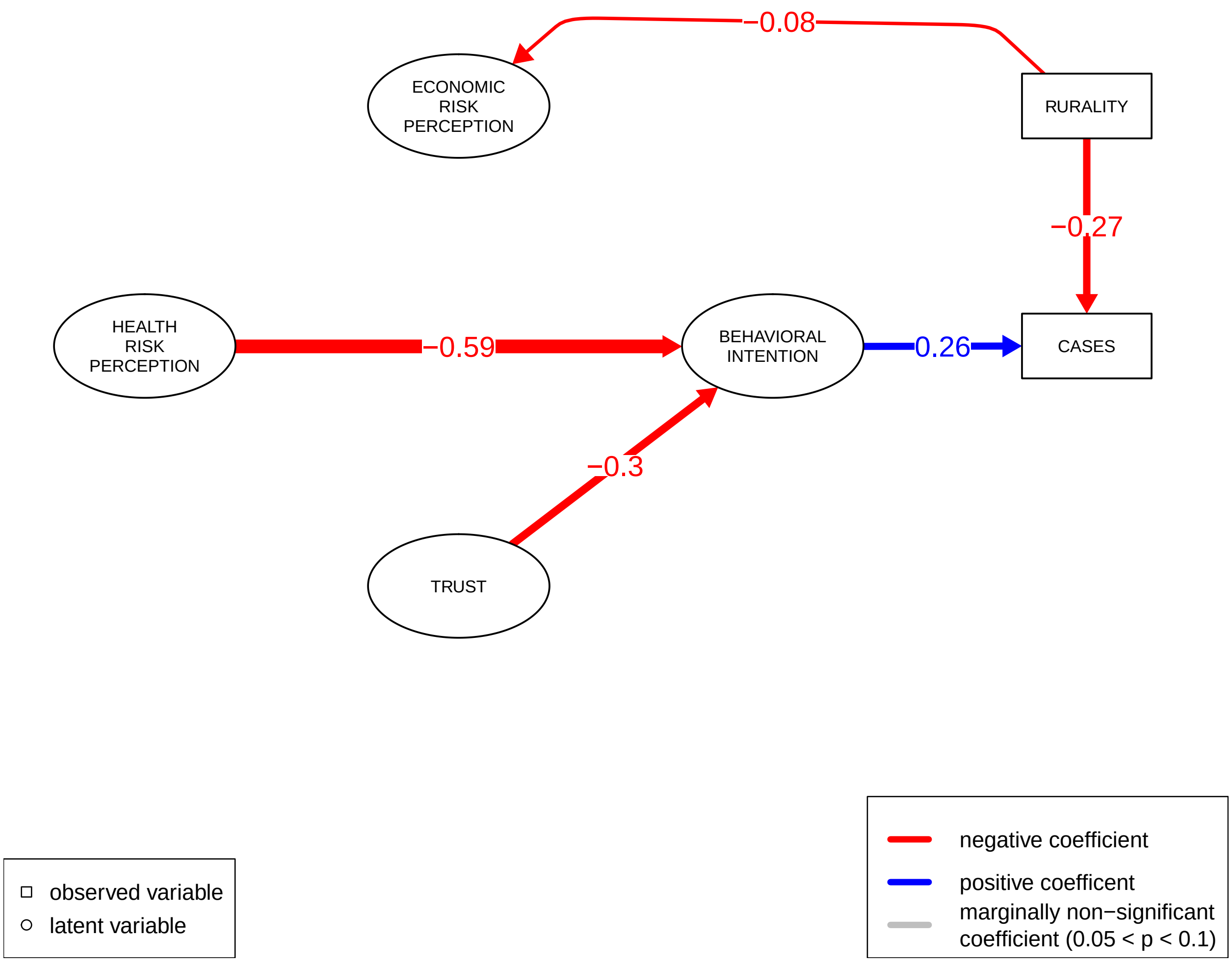

#### Table S3.6: Model 1B - Trust to Risk Perception on Behavioral Intention Summary Table Output

| Latent Variable | Regression Variable | std.est | std.error | z | p-value |
|---|---|---|---|---|---|
| beh_int | indoor_close | 0.6318594 | 0.000 | NA | NA |
| | indoor_group | 0.8600134 | 1.245 | 2.399 | 0.016 |
| | restaurant | 0.8625935 | 0.892 | 2.149 | 0.032 |
| | church | 0.8194615 | 2.645 | 2.582 | 0.010 |
| | shop | 0.4832687 | 0.395 | 1.696 | 0.090 |
| | appointments | 0.6340223 | 0.529 | 1.771 | 0.077 |
| | community | 0.8597569 | 0.906 | 2.180 | 0.029 |
| | mask | 0.6159526 | 3.233 | 2.595 | 0.009 |
| trust | ruralbool | -0.0524177 | 0.027 | -1.709 | 0.088 |
| | femalebool | 0.0471707 | 0.027 | 1.529 | 0.126 |
| | whitebool | 0.1198171 | 0.047 | 3.744 | 0.000 |
| | age65plusbool | 0.0355333 | 0.029 | 1.152 | 0.250 |
| | polVecLiberal | 0.1269346 | 0.036 | 3.468 | 0.001 |
| | polVecConservative | -0.4792851 | 0.049 | -9.542 | 0.000 |
| | polVecLibertarian | -0.2718985 | 0.087 | -7.262 | 0.000 |
| | polVecNonpolitical | -0.1254447 | 0.057 | -3.671 | 0.000 |
| | polVecOther | -0.0278327 | 0.099 | -0.898 | 0.369 |
| health_risk | trust | 0.5973360 | 0.112 | 9.939 | 0.000 |
| | ruralbool | 0.0174238 | 0.047 | 0.622 | 0.534 |
| | femalebool | 0.1073022 | 0.047 | 3.791 | 0.000 |
| | whitebool | -0.1218331 | 0.078 | -4.238 | 0.000 |
| | age65plusbool | 0.2030798 | 0.050 | 7.091 | 0.000 |
| | polVecLiberal | 0.0818772 | 0.060 | 2.488 | 0.013 |
| | polVecConservative | -0.0504230 | 0.069 | -1.341 | 0.180 |
| | polVecLibertarian | -0.0606063 | 0.134 | -1.962 | 0.050 |
| | polVecNonpolitical | 0.0273616 | 0.096 | 0.894 | 0.371 |
| | polVecOther | 0.0432130 | 0.170 | 1.520 | 0.129 |
| econ_risk | trust | 0.2290899 | 0.052 | 3.819 | 0.000 |
| | ruralbool | -0.0656218 | 0.030 | -1.672 | 0.094 |
| | femalebool | 0.0837533 | 0.030 | 2.103 | 0.035 |
| | whitebool | -0.0643906 | 0.051 | -1.609 | 0.108 |
| | age65plusbool | 0.0035750 | 0.032 | 0.092 | 0.927 |
| | polVecLiberal | 0.0507565 | 0.039 | 1.112 | 0.266 |
| | polVecConservative | -0.1511760 | 0.046 | -2.804 | 0.005 |
| | polVecLibertarian | 0.0995460 | 0.088 | 2.278 | 0.023 |
| | polVecNonpolitical | -0.0080773 | 0.062 | -0.191 | 0.848 |
| | polVecOther | 0.0210756 | 0.109 | 0.536 | 0.592 |
| beh_int | econ_risk | 0.0709011 | 0.006 | 1.790 | 0.073 |
| | health_risk | -0.7169805 | 0.021 | -2.602 | 0.009 |
| | ruralbool | 0.0380051 | 0.003 | 1.372 | 0.170 |
| | femalebool | 0.0649470 | 0.004 | 1.905 | 0.057 |
| | whitebool | -0.0308915 | 0.005 | -1.191 | 0.234 |
| | age65plusbool | 0.0007897 | 0.003 | 0.033 | 0.974 |
| | polVecLiberal | -0.0563036 | 0.005 | -1.613 | 0.107 |
| | polVecConservative | 0.2140480 | 0.012 | 2.469 | 0.014 |
| | polVecLibertarian | 0.0564483 | 0.011 | 1.726 | 0.084 |
| | polVecNonpolitical | 0.0281905 | 0.006 | 1.047 | 0.295 |
| | polVecOther | 0.0121386 | 0.010 | 0.514 | 0.607 |
| | casetodateper100 | 0.1280073 | 0.002 | 2.351 | 0.019 |

*Note:*

chisq = 10336.253 aic = 89908.147 rmsea = 0.071

#### Figure S3.10: Model 1B - Trust to Risk Perception on Behavioral Intention Full Diagram

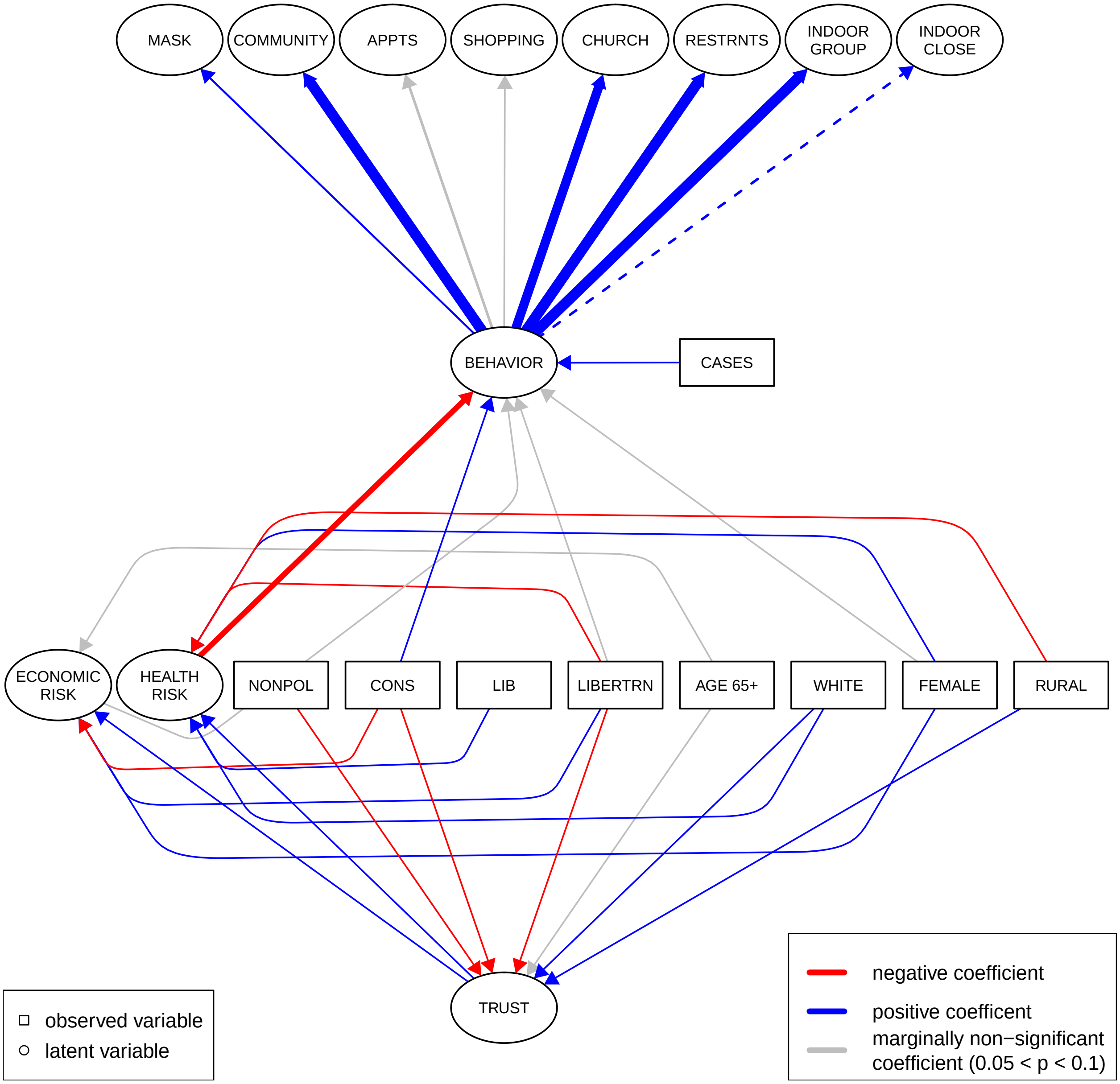

#### Figure S3.11: Model 1B - Trust to Risk Perception on Behavioral Intention Simplified Diagram

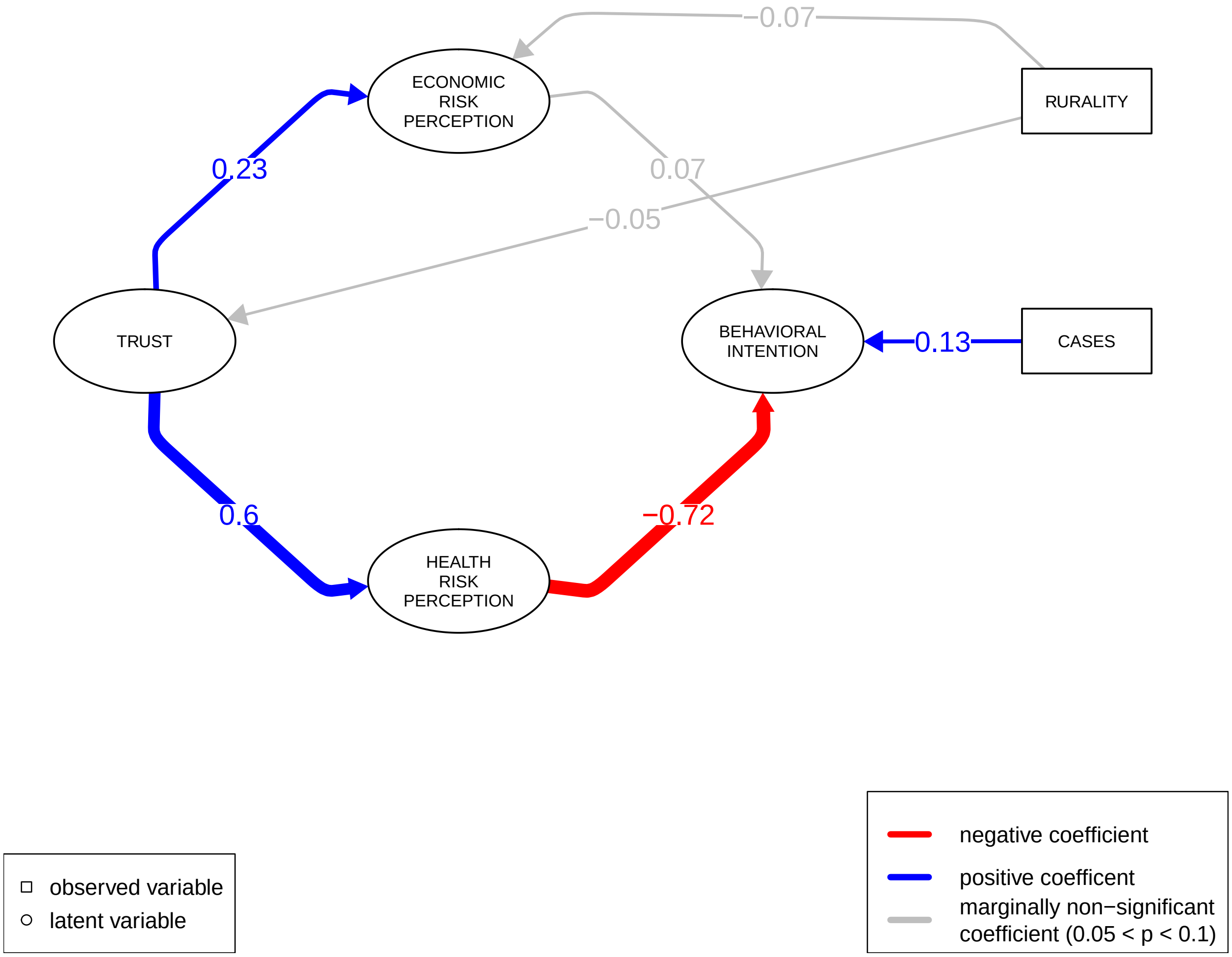

#### Table S3.7: Model 2B - Risk Perception to Trust on Behavioral Intention Summary Table Ouput

| Latent Variable | Regression Variable | std.est | std.error | z | p-value |
|---|---|---|---|---|---|
| beh__int | indoor__close | 0.6126064 | 0.000 | NA | NA |
| | indoor__group | 0.8656567 | 1.276 | 2.403 | 0.016 |
| | restaurant | 0.8556833 | 0.905 | 2.151 | 0.031 |
| | church | 0.8167390 | 2.677 | 2.589 | 0.010 |
| | shop | 0.4691448 | 0.395 | 1.703 | 0.089 |
| | appointments | 0.6267748 | 0.539 | 1.781 | 0.075 |
| | community | 0.8611842 | 0.926 | 2.181 | 0.029 |
| | mask | 0.6166035 | 3.299 | 2.603 | 0.009 |
| trust | ruralbool | -0.0463969 | 0.023 | -1.745 | 0.081 |
| | femalebool | -0.0271250 | 0.023 | -1.003 | 0.316 |
| | whitebool | 0.1469592 | 0.041 | 5.101 | 0.000 |
| | age65plusbool | -0.0903822 | 0.026 | -3.187 | 0.001 |
| | polVecLiberal | 0.0397291 | 0.030 | 1.274 | 0.203 |
| | polVecConservative | -0.2980511 | 0.039 | -7.363 | 0.000 |
| | polVecLibertarian | -0.1450684 | 0.069 | -4.785 | 0.000 |
| | polVecNonpolitical | -0.0991899 | 0.048 | -3.374 | 0.001 |
| | polVecOther | -0.0420054 | 0.084 | -1.564 | 0.118 |
| | health__risk | 0.5672902 | 0.029 | 10.167 | 0.000 |
| | econ__risk | -0.0285050 | 0.037 | -0.903 | 0.367 |
| health__risk | ruralbool | -0.0145495 | 0.052 | -0.461 | 0.645 |
| | femalebool | 0.1362395 | 0.052 | 4.264 | 0.000 |
| | whitebool | -0.0498488 | 0.087 | -1.561 | 0.118 |
| | age65plusbool | 0.2228096 | 0.056 | 6.895 | 0.000 |
| | polVecLiberal | 0.1577046 | 0.067 | 4.276 | 0.000 |
| | polVecConservative | -0.3373909 | 0.069 | -8.933 | 0.000 |
| | polVecLibertarian | -0.2231377 | 0.143 | -6.724 | 0.000 |
| | polVecNonpolitical | -0.0483584 | 0.107 | -1.417 | 0.157 |
| | polVecOther | 0.0255500 | 0.191 | 0.796 | 0.426 |
| econ__risk | ruralbool | -0.0788062 | 0.029 | -2.004 | 0.045 |
| | femalebool | 0.0915249 | 0.030 | 2.295 | 0.022 |
| | whitebool | -0.0353921 | 0.048 | -0.906 | 0.365 |
| | age65plusbool | 0.0088043 | 0.031 | 0.226 | 0.821 |
| | polVecLiberal | 0.0775047 | 0.037 | 1.706 | 0.088 |
| | polVecConservative | -0.2532709 | 0.043 | -4.860 | 0.000 |
| | polVecLibertarian | 0.0439062 | 0.078 | 1.092 | 0.275 |
| | polVecNonpolitical | -0.0378203 | 0.059 | -0.906 | 0.365 |
| | polVecOther | 0.0133595 | 0.105 | 0.342 | 0.733 |
| beh__int | trust | -0.6204804 | 0.034 | -2.552 | 0.011 |
| | ruralbool | -0.0123779 | 0.003 | -0.456 | 0.648 |
| | femalebool | 0.0097836 | 0.003 | 0.371 | 0.711 |
| | whitebool | 0.0728175 | 0.008 | 1.900 | 0.057 |
| | age65plusbool | -0.1450171 | 0.008 | -2.376 | 0.018 |
| | polVecLiberal | -0.1073202 | 0.007 | -2.093 | 0.036 |
| | polVecConservative | 0.1544940 | 0.009 | 2.259 | 0.024 |
| | polVecLibertarian | 0.0533238 | 0.011 | 1.522 | 0.128 |
| | polVecNonpolitical | -0.0185510 | 0.007 | -0.636 | 0.525 |
| | polVecOther | -0.0236812 | 0.012 | -0.852 | 0.394 |
| | casetodateper100 | 0.0304744 | 0.001 | 1.025 | 0.305 |

*Note:*

chisq = 10552.368 aic = 90122.262 rmsea = 0.072

#### Figure S3.14: Model 2B - Risk Perception to Trust on Behavioral Intention Full Diagram

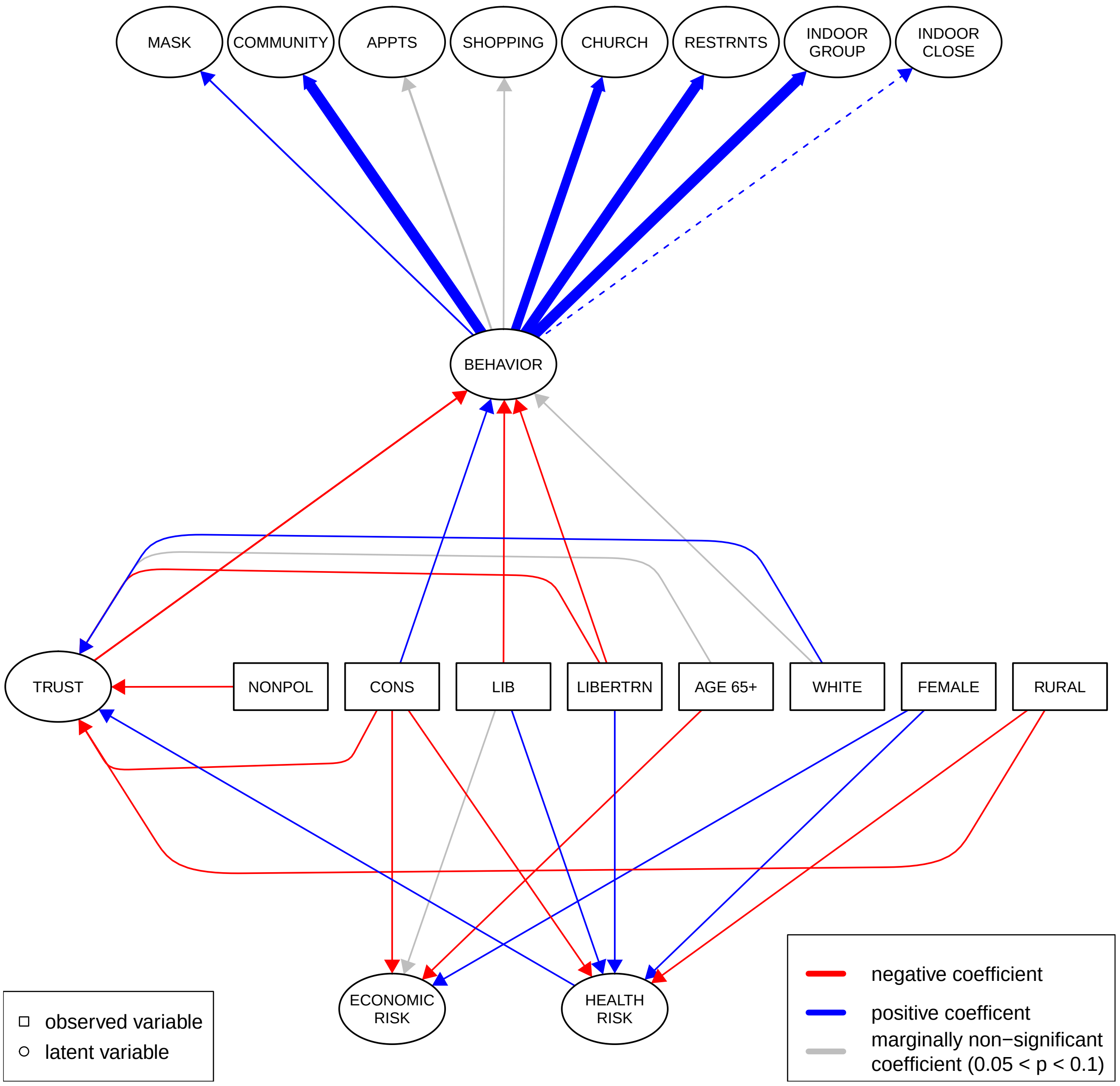

#### Figure S3.15: Model 2B - Risk Perception to Trust on Behavioral Intention Simplified Diagram

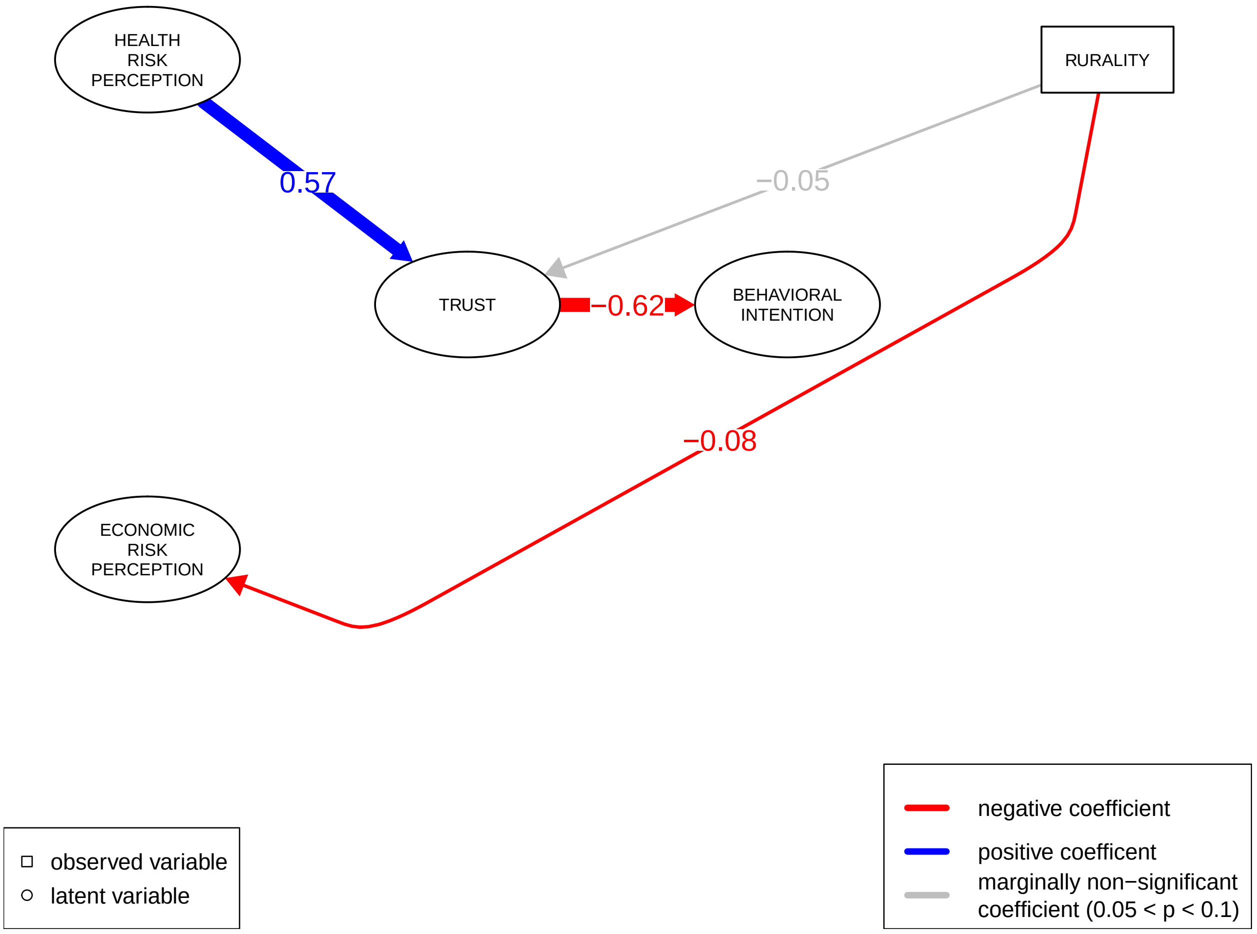

#### Table S3.8: Model 3B - Trust and Risk Perception on Behavioral Intention Summary Table Output

| Latent Variable | Regression Variable | std.est | std.error | z | p-value |
|---|---|---|---|---|---|
| beh_int | indoor_close | 0.6144116 | 0.000 | NA | NA |
| | indoor_group | 0.8546174 | 1.274 | 2.356 | 0.018 |
| | restaurant | 0.8501598 | 0.909 | 2.107 | 0.035 |
| | church | 0.8081635 | 2.688 | 2.542 | 0.011 |
| | shop | 0.4638008 | 0.397 | 1.675 | 0.094 |
| | appointments | 0.6154355 | 0.536 | 1.742 | 0.081 |
| | community | 0.8519501 | 0.927 | 2.137 | 0.033 |
| | mask | 0.6071950 | 3.321 | 2.556 | 0.011 |
| trust | ruralbool | -0.0507530 | 0.029 | -1.645 | 0.100 |
| | femalebool | 0.0453215 | 0.029 | 1.461 | 0.144 |
| | whitebool | 0.1232736 | 0.050 | 3.841 | 0.000 |
| | age65plusbool | 0.0377615 | 0.031 | 1.215 | 0.224 |
| | polVecLiberal | 0.1262213 | 0.038 | 3.443 | 0.001 |
| | polVecConservative | -0.4713280 | 0.050 | -9.791 | 0.000 |
| | polVecLibertarian | -0.2697549 | 0.091 | -7.340 | 0.000 |
| | polVecNonpolitical | -0.1250477 | 0.062 | -3.656 | 0.000 |
| | polVecOther | -0.0292698 | 0.107 | -0.938 | 0.348 |
| health_risk | ruralbool | -0.0129635 | 0.054 | -0.410 | 0.682 |
| | femalebool | 0.1336858 | 0.054 | 4.180 | 0.000 |
| | whitebool | -0.0508124 | 0.089 | -1.588 | 0.112 |
| | age65plusbool | 0.2272247 | 0.058 | 7.029 | 0.000 |
| | polVecLiberal | 0.1561511 | 0.068 | 4.229 | 0.000 |
| | polVecConservative | -0.3315642 | 0.070 | -8.796 | 0.000 |
| | polVecLibertarian | -0.2202329 | 0.146 | -6.637 | 0.000 |
| | polVecNonpolitical | -0.0451669 | 0.109 | -1.321 | 0.187 |
| | polVecOther | 0.0290602 | 0.195 | 0.904 | 0.366 |
| econ_risk | ruralbool | -0.0789920 | 0.029 | -2.010 | 0.044 |
| | femalebool | 0.0912450 | 0.029 | 2.290 | 0.022 |
| | whitebool | -0.0351966 | 0.047 | -0.903 | 0.367 |
| | age65plusbool | 0.0086715 | 0.030 | 0.223 | 0.823 |
| | polVecLiberal | 0.0772715 | 0.037 | 1.703 | 0.089 |
| | polVecConservative | -0.2524772 | 0.042 | -4.845 | 0.000 |
| | polVecLibertarian | 0.0444537 | 0.078 | 1.107 | 0.268 |
| | polVecNonpolitical | -0.0379610 | 0.059 | -0.910 | 0.363 |
| | polVecOther | 0.0131611 | 0.104 | 0.337 | 0.736 |
| beh_int | econ_risk | 0.0680950 | 0.006 | 1.747 | 0.081 |
| | health_risk | -0.6193686 | 0.017 | -2.559 | 0.010 |
| | trust | -0.2577844 | 0.013 | -2.428 | 0.015 |
| | ruralbool | 0.0220278 | 0.003 | 0.862 | 0.389 |
| | femalebool | 0.0633930 | 0.004 | 1.850 | 0.064 |
| | whitebool | 0.0039178 | 0.005 | 0.164 | 0.870 |
| | age65plusbool | -0.0178966 | 0.003 | -0.704 | 0.481 |
| | polVecLiberal | -0.0518845 | 0.005 | -1.494 | 0.135 |
| | polVecConservative | 0.1485000 | 0.009 | 2.233 | 0.026 |
| | polVecLibertarian | 0.0194691 | 0.008 | 0.709 | 0.478 |
| | polVecNonpolitical | 0.0041008 | 0.006 | 0.161 | 0.872 |
| | polVecOther | 0.0034422 | 0.010 | 0.146 | 0.884 |
| | casetodateper100 | 0.1044174 | 0.002 | 2.205 | 0.027 |

*Note:*

chisq = 10509.767 aic = 90079.661 rmsea = 0.072

#### Figure S3.12: Model 3B - Trust and Risk Perception on Behavioral Intention Full Diagram

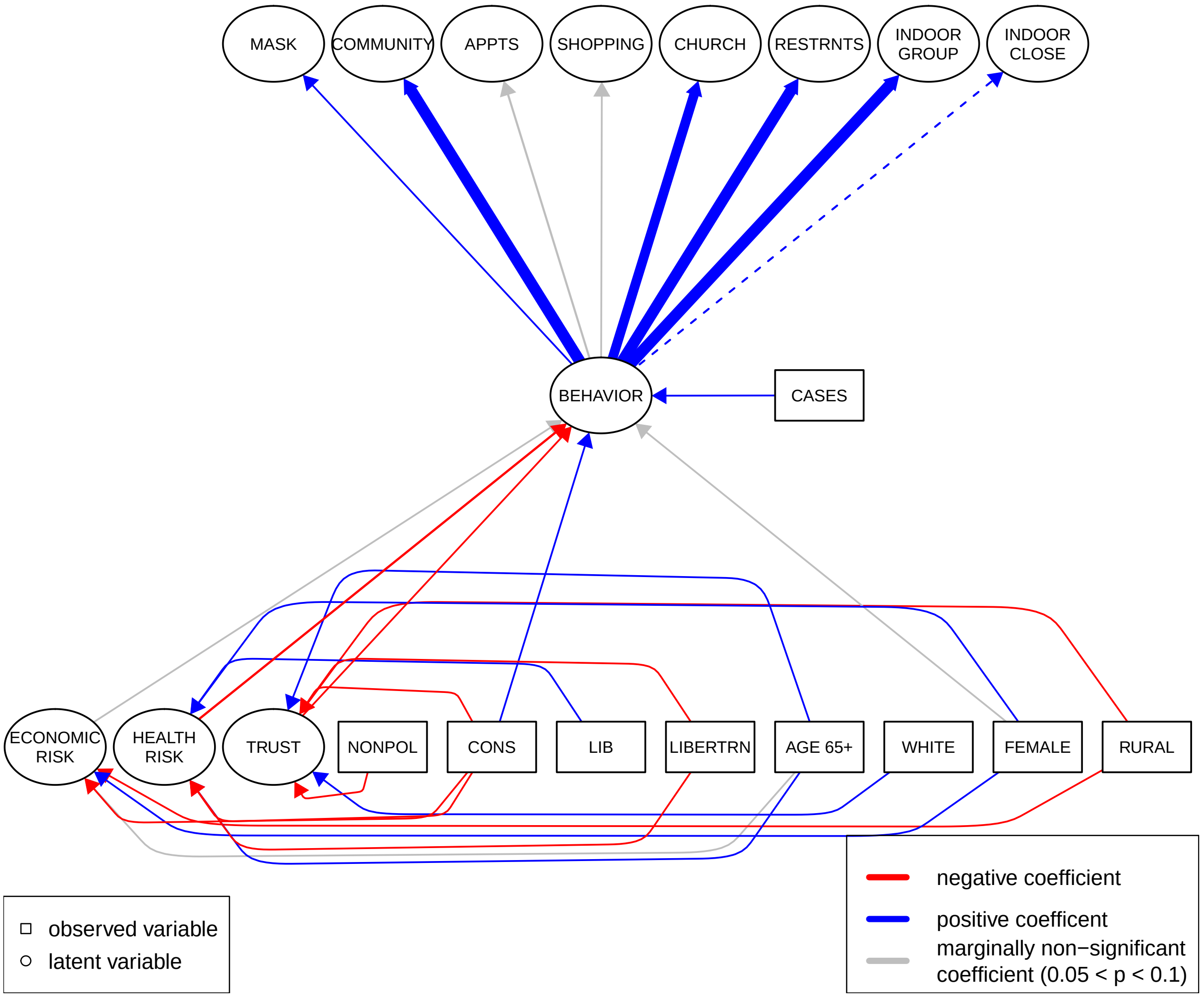

#### Figure S3.13: Model 3B - Trust and Risk Perception on Behavioral Intention Simplified Diagram

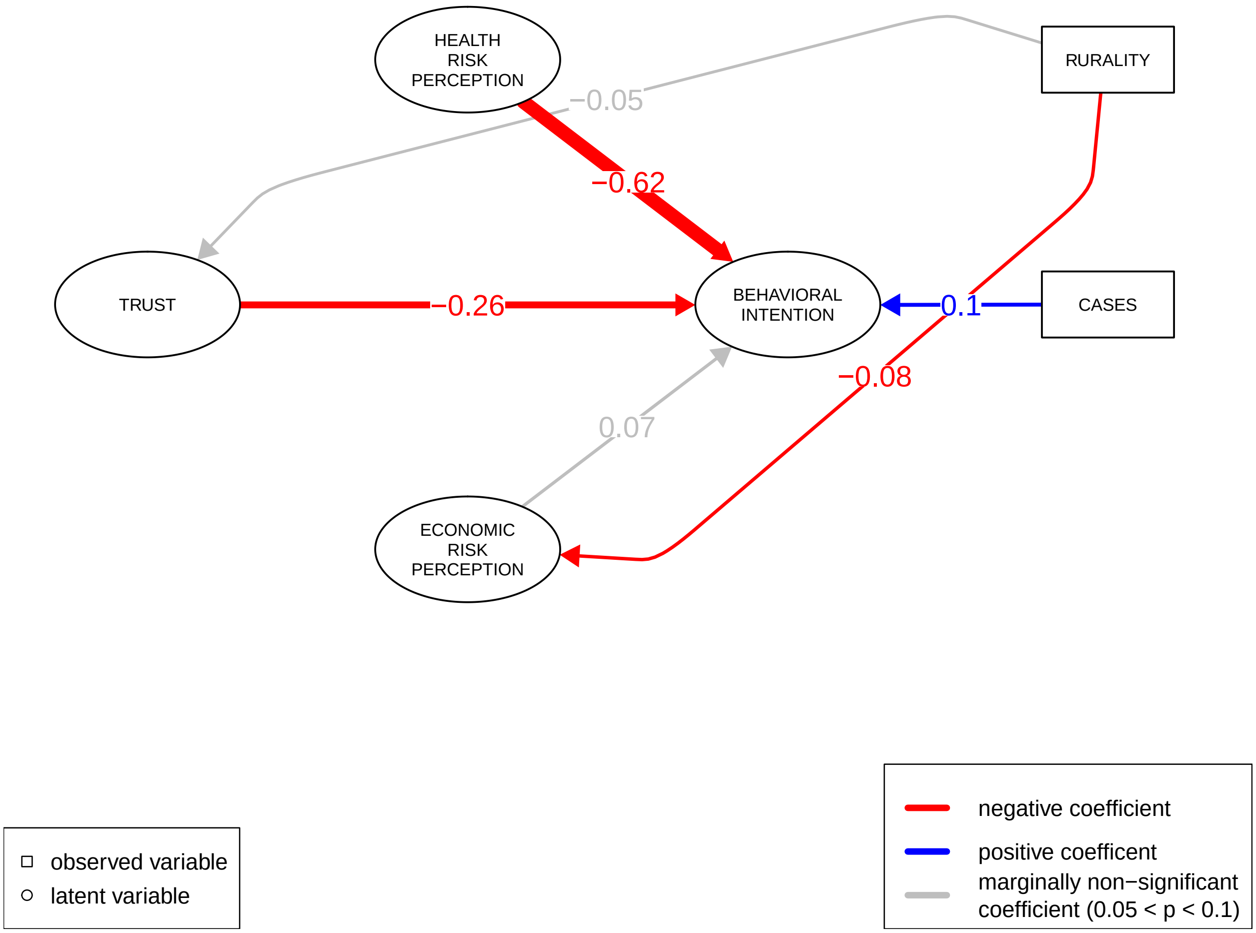

##### Table S3.9: Comparing Model Set A

```
## Chi-Squared Difference Test
##
##           Df   AIC   BIC Chisq Chisq diff Df diff Pr(>Chisq)
## model1A 2003 98774 99543 10470
## model2A 2004 98950 99715 10648     178.41       1  < 2.2e-16 ***
## ---
## Signif. codes:  0 '***' 0.001 '**' 0.01 '*' 0.05 '.' 0.1 ' ' 1
```

```
## Chi-Squared Difference Test
##
##           Df   AIC   BIC Chisq Chisq diff Df diff Pr(>Chisq)
## model1A 2003 98774 99543 10470
## model3A 2004 98932 99697 10630     159.91       1  < 2.2e-16 ***
## ---
## Signif. codes:  0 '***' 0.001 '**' 0.01 '*' 0.05 '.' 0.1 ' ' 1
```

#### Table S3.10: Comparing Model Set B

```
## Chi-Squared Difference Test
##
##           Df   AIC   BIC Chisq Chisq diff Df diff Pr(>Chisq)
## model1B 1995 89908 90668 10336
## model2B 1996 90122 90877 10552     216.12       1  < 2.2e-16 ***
## ---
## Signif. codes:  0 '***' 0.001 '**' 0.01 '*' 0.05 '.' 0.1 ' ' 1
```

```
## Chi-Squared Difference Test
##
##           Df   AIC   BIC Chisq Chisq diff Df diff Pr(>Chisq)
## model1B 1995 89908 90668 10336
## model3B 1996 90080 90835 10510     173.51       1  < 2.2e-16 ***
## ---
## Signif. codes:  0 '***' 0.001 '**' 0.01 '*' 0.05 '.' 0.1 ' ' 1
```
